# HEALTHCARE UTILISATION IN PATIENTS WITH LONG-TERM CONDITIONS DURING THE COVID-19 PANDEMIC: A POPULATION BASED STUDY ACROSS GREATER MANCHESTER, UK

**DOI:** 10.1101/2022.06.09.22276232

**Authors:** Camilla Sammut-Powell, Richard Williams, Matthew Sperrin, Owain Thomas, Niels Peek, Stuart W. Grant

## Abstract

**Background:** The COVID-19 pandemic has placed an unprecedented demand on global healthcare resources. Data on whole population healthcare utilisation (HCU) across both primary and secondary care during the COVID-19 pandemic are lacking.

**Aim:** To describe primary and secondary HCU stratified by LTCs and deprivation, during the first 19 months of COVID-19 pandemic across a large urban area in the United Kingdom.

**Methods:** Observational HCU data between 30^th^-December-2019 and 1^st^-August-2021 were extracted from the Greater Manchester Care Record. Primary care HCU (incident prescribing and recording of healthcare information) and secondary care HCU (planned and unplanned admissions) were assessed.

**Results:** The first national lockdown was associated with reductions in all primary HCU measures, ranging from 24.7% (24.0% to 25.5%) for incident prescribing to 84.9% (84.2% to 85.5%) for cholesterol monitoring. Secondary HCU also dropped significantly for planned (47% (42.9% to 51.5%)) and unplanned admissions (35.0% (28.3% to 41.6%)). Only secondary care had significant reductions in HCU during the second national lockdown. Primary HCU measures had not recovered to pre-pandemic levels by the end of the study. The secondary admission rate ratio between multi-morbid patients and those without LTCs increased during the first lockdown by a factor of 2.4 (2.0 to 2.9;p<0.001) for planned admissions and by 1.3 (1.1 to 1.5;p=0.006) for unplanned admissions. No significant changes in this ratio were observed in primary HCU. Different patterns in secondary care HCU were observed by LTC group.

**Conclusion:** Major changes in primary and secondary HCU have been observed during the COVID-19 pandemic. Secondary HCU reduced more in those without LTCs and the ratio of utilisation between the most and least deprived increased for the majority of HCU measures. Overall primary HCU measures and secondary care HCU for some LTC groups had not returned to pre-pandemic levels by the end of the study.

## INTRODUCTION

On the 30th of January 2020, the World Health Organization (WHO) declared a public health emergency of international concern with governments urged to prepare for global spread of COVID-19.^1^ With case numbers increasing and the virus spreading globally, COVID-19 was characterised as a pandemic six weeks later and rapidly developed into a global public health emergency. As of the 17^th^ of December 2021 approximately 273 million cases and 5.3 million COVID-19 associated deaths have been reported globally.^2^Governments across the world enacted a range of measures aimed at controlling the spread of the virus,^3^ and increasing healthcare capacity.^4,5^ Despite these measures healthcare systems have been overwhelmed and diversion of healthcare resources to address increased demand specific to COVID-19 has been required.^6,7^ The impact of this diversion of resources on the care of patients with non COVID-19 illnesses has been exacerbated by reduced staff availability due to COVID-19 infection amongst healthcare workers.^8^

Numerous studies have been undertaken to assess the impact of the pandemic on healthcare provision in a variety of settings. An analysis of UK general practitioner (GP) data demonstrated that diagnoses of common physical and mental health conditions decreased substantially early in the pandemic.^9^ The number of urgent GP referrals for cancer fell by 60% in April 2020 compared to the same month in 2019.^10^ Hospital administrative data has demonstrated a decline in patients presenting with acute coronary syndrome from mid-February 2020 onwards, ^11^ and a separate analysis demonstrated a 43% reduction in patients undergoing percutaneous coronary interventions for ST-elevation myocardial infarctions compared to previous years.^12^ Modelling studies have suggested that approximately 28,000,000 elective surgical procedures were cancelled over a 12-week period of peak disruption caused by the pandemic.^13^

Most studies to date investigating the impact of the pandemic on healthcare utilisation (HCU) have assessed specific patient groups, largely focussed on secondary care.^14^ Changes to HCU during the COVID-19 pandemic for both primary and secondary care stratified across the range of long-term medical conditions (LTCs) and different levels of social deprivation have not previously been described. The Greater Manchester Care Record (GMCR) includes electronic health records from all primary and secondary care National Health Service (NHS) providers in the metropolitan county of Greater Manchester (GM). GM has been significantly affected by COVID-19,^15^ and the GMCR provides a unique opportunity to study the impact of the pandemic on primary and secondary HCU in patients with LTCs across this defined urban area.

## METHODS

### Design and Data Source

This was a retrospective, observational, service evaluation using routinely collected data. The data analysed were from two sources: 1) HCU data from the GMCR which is an integrated patient record containing data from primary and secondary NHS services across Greater Manchester (GM) and 2) contextual Government COVID-19 data ^16^ regarding the number of new COVID-19 cases and COVID-19 related hospital admissions.

### Greater Manchester Care Record

The GMCR is populated with data from primary care (General Practitioners), secondary care (acute and community hospitals), mental health trusts and social care across an entire geographical region. A total of nine secondary care organisations (including 12 hospitals), 3 mental health trusts and 10 clinical commissioning groups (CCGs) contribute data. The primary purpose of the GMCR is for direct patient care as it provides clinicians with information from other health care providers relevant to their patient encounters that would ordinarily be inaccessible. However, it has also been made available in de-identified format for research relating to COVID-19.

### UK Government COVID-19 Data

Data regarding the number of new COVID-19 cases and COVID-19 related hospital admissions were collected by the UK government throughout the pandemic. The number of new cases by specimen date was extracted for Manchester and the number of COVID-19 admissions were extracted for each of the secondary acute providers serving the people with a Manchester CCG (MCCG), included within the GMCR. The data is freely available from https://coronavirus.data.gov.uk/details/download and full details of the data extraction are provided in the Supplementary Materials.

### Data processing and approvals

All identifiable data including free text are redacted. Some non-identifying demographic data are available such as recorded gender, year of birth, lower layer super output area (LSOA), index of multiple deprivation and ethnicity. The University of Manchester is permitted to perform research on this data via a Greater Manchester wide data protection impact assessment (DPIA). The basis for this DPIA is the control of patient information (COPI) notice issued by the Secretary of State for Health and Social Care in March 2020 which allowed confidential patient information to be shared for the purposes of research into COVID-19.^17^ The study was approved by both the GMCR Expert Review Group and Research Governance Group. All data made available to the analysts was de-identified and aggregated and therefore did not require specific ethical approval.

### Study populations and key time points

The main study population consisted of all patients that were registered with a GP within GM on 1^st^ January 2020, defined as the GM population. The 1^st^ January 2020 is the index study date. For the primary care analyses the entire GM population were considered. However, secondary care data were only available for patients registered to a Manchester CCG, hence, the secondary healthcare utilisation analyses were limited to these people. The dates of the national lockdowns initiated in response to the COVID-19 pandemic were indicated in addition to Christmas week due to expected changes in HCU during these periods. The first national lockdown ran from the 23^rd^ March 2020 to the 11^th^ May 2020, the second national lockdown ran from 5^th^ November 2020 to 1^st^ December 2020 and the third national lockdown ran from 6^th^ January 2021 to the 8^th^ March 2021.

### Long-term medical conditions

Long-term medical conditions (LTCs) were defined as per Barnett et al,^18^ and were grouped into the following categories: cancer, cardiovascular, endocrine, gastrointestinal, musculoskeletal or skin, neurological, psychiatric, renal or urological, respiratory, sensory impairment or learning disability, and substance abuse. A resident was identified as being diagnosed with a LTC by interrogating the GMCR record prior to the index date. If a long-term medical condition was diagnosed after the 1^st^ January 2020 the patient was not recorded as having the LTC for this analysis. People that were identified as belonging to multiple LTC groups were assigned to each corresponding LTC group and defined as multi-morbid. The full list of long-term conditions and groupings are provided in Supplementary Table S1.

### Index of Multiple Deprivation

The 2019 Index of Multiple Deprivation (IMD) is the official measure of relative deprivation provided by the Office for National Statistics which combines information from seven different domains to produce an overall relative measure of deprivation for each LSOA. Each LSOA is ranked from least to most deprived, and deciles of relative deprivation are generated.^19^ For this study the available IMD deciles were categorised into four groups, representing the most deprived (deciles 1-2), highly deprived (deciles 3-4), moderately deprived (deciles 5-6) and the least deprived LSOAs (deciles 7-10). The least deprived group consisted of 4 deciles to avoid multiple small groups because of the skew towards more deprived deciles in GM.

### Measuring Healthcare Utilisation

For primary HCU, surrogate markers were evaluated consisting of first prescriptions and recording of healthcare information in the GP record. First prescriptions were identified by the issuing of any new prescription for an individual patient by a primary care healthcare professional and are referred to as incident prescriptions throughout the manuscript. This measure was selected as issuing of an incident prescription requires contact with a healthcare professional. Healthcare information recorded included: recoding of smoking status, measurements of cholesterol, blood pressure (BP), blood glucose (HbA1c) and body mass index (BMI). The values of these measurements were not used in the analysis.

For secondary HCU, the number of acute provider admissions were evaluated. To enable population admission rates to be calculated, a denominator was calculated by assigning each resident within Manchester CCG to a secondary care provider according to the most common provider observed within their LSOA. In cases where there the most common secondary provider was unclear one of the two most common providers was randomly assigned to that LSOA. Secondary care admissions were categorised into planned, unplanned, maternity, transfers and ‘other’ admission, defined according to the admission type field available in the provider data. Daily aggregate level data counts of all utilisation measures were provided. A full description of the data processing applied is available at https://github.com/rw251/gm-idcr/tree/master/projects/001%20-%20Grant.

### Statistical Analysis

Weekly totals of HCU data were evaluated for the entire population. The rate ratios (RRs) of utilisation in the weeks before and after the initiation of the first and second national lockdowns (1^st^ national lockdown: w/c 23^rd^ March 2020 vs w/c 9^th^ March 2020; 2^nd^ national lockdown: w/c 9^th^ November 2020 vs w/c 19^th^ October 2020) were estimated across all measures of HCU to determine the association between the initiation of each national lockdown and HCU.

The effect of the initiation of the third national lockdown was not estimated since the weeks prior coincided with Christmas, where utilisation is expected to be reduced. The pre-pandemic weeks (prior to 9^th^ March 2020) were compared against each of the national lockdown periods to determine if there was a significant change in the rates of HCU associated with each of the national lockdowns using Poisson regression. A Poisson regression model, linear in time, was fit to the weekly rates of utilisation after the initiation of the first national lockdown until the end of the study to determine the overall change of utilisation throughout the pandemic. A direct comparison between utilisation observed in a calendar week in 2021 vs 2020 was conducted for calendar weeks 2 to 11, using rate ratios; it was assumed that the data in calendar weeks 2 to 11 in 2020 were unaffected by the pandemic and consequently act as a control. Additionally, the rate ratios between utilisation measures in the final four weeks of the study and the pre-pandemic period were estimated using Poisson regression to compare how utilisation differed from pre-pandemic levels by the end of the study.

Subgroup analyses were performed across LTC and IMD groups. To compare subgroups, we further provided the rates of utilisation per 1000 people by dividing by the total number of people assigned to the corresponding subgroup and multiplying by 1000. For example, when comparing the rates across number of LTCs (none, one or multiple), for the people without any LTCs, the rate of weekly secondary care admissions is defined as the total number of admissions experienced by this subgroup within a given week divided by the total number of people within the subgroup, multiplied by 1000. The interactive effect between the each of the national lockdowns and subgroup HCU was estimated using log-linear regression.

A sensitivity analysis to adjust for deaths that occurred during the study was performed, where rates of utilisation were re-calculated in July 2021 dividing by the total numbers of patients that were still alive (death-adjusted), and compared to the un-adjusted rates. July 2021 was chosen since this was one month before the end of the study and therefore captured the majority of deaths that occurred throughout the study; hence if no difference was observed between the death-adjusted and un-adjusted rates, the un-adjusted rates would pertain across all weeks. All analyses were performed in R version 4.0.0, using the packages ‘tidyverse’^20^, ‘scales’^21^, ‘reshape2’^22^ and ‘cowplot’^23^.

### Patient and public involvement

Two public representatives provided input throughout the project. Both representatives gave their full support to the proposed project and are preparing a patient and public summary of the research for dissemination.

## RESULTS

The total population captured within the GMCR includes 3,225,169 patients, of whom 693,749 were registered with a Manchester CCG. The prevalence of LTCs is shown in Table 1. The most common LTCs observed were psychiatric (GMCR: 26.5%, MCCG: 20.7%), cardiovascular (GMCR: 17.4%, MCCG: 10.5%), respiratory (GMCR: 17.1%, MCCG: 13.4%) and gastrointestinal (GMCR: 15.5%, MCCG: 11.8%). Levels of deprivation were high, with 41.4% of the GM population and the majority of those registered within Manchester CCG (58.5%) in the most deprived quintile (decile of 1 or 2; Table 1).

**Table 1:** Long-term conditions and social deprivation identified in the GM population and the Manchester CCG subpopulation.

|  | Greater Manchester<br>(N=3,225,169) | % | Manchester CCG<br>(N=693,749) | % |
| --- | --- | --- | --- | --- |
| **LTC Group\*** |  |  |  |  |
| Cancer | 50954 | 1.6 | 6307 | 0.9 |
| Cardiovascular | 561195 | 17.4 | 72912 | 10.5 |
| Endocrine | 393274 | 12.2 | 64557 | 9.3 |
| Gastrointestinal | 501060 | 15.5 | 81957 | 11.8 |
| Musculoskeletal or Skin | 284103 | 8.8 | 52593 | 7.6 |
| Neurological | 46672 | 1.4 | 7340 | 1.1 |
| Psychiatric | 854454 | 26.5 | 143707 | 20.7 |
| Renal or Urological | 130436 | 4.0 | 16617 | 2.4 |
| Respiratory | 550648 | 17.1 | 93174 | 13.4 |
| Sensory Impairment or Learning Disability | 314264 | 9.7 | 45909 | 6.6 |
| Substance Abuse | 115532 | 3.6 | 24068 | 3.5 |
| **Number of LTCs** |  |  |  |  |
| None | 1530501 | 47.5 | 399618 | 57.6 |
| One | 631648 | 19.6 | 127428 | 18.4 |
| Multiple | 1063020 | 33.0 | 166703 | 24.0 |
| **IMD Group** |  |  |  |  |
| 1 to 2 | 1335061 | 41.4 | 405862 | 58.5 |
| 3 to 4 | 675296 | 20.9 | 185118 | 26.7 |
| 5 to 6 | 424139 | 13.2 | 67192 | 9.7 |
| 7 to 10 | 788553 | 24.4 | 35087 | 5.1 |
| Missing | 2120 | 0.1 | 490 | 0.1 |

LTC – long term condition; IMD – index of multiple deprivation. \*These data represent the overall prevalence of each long term condition in the population. Each individual can be represented in more than one LTC row.

### Overall primary HCU

There was a rapid decrease in all weekly primary HCU measures starting just prior to the first national lockdown (Figure 1). The largest drops in activity associated with the initiation of the first national lockdown were for recording of healthcare information (% drop (95% CI): BP: 82.4% (82.0%-82.9%); BMI: 79.5% (78.8%-80.1%); Cholesterol 84.9% (84.2%-85.5%); HbA1c 84.0% (83.3%-84.6%); Smoking status 62.2% (61.3%-63.1%). There was still a significant drop in the new prescriptions but the change was proportionally smaller (24.7%; 95% CI: 24.0%-25.5%). These reductions were sustained throughout the first national lockdown (Supplementary Table S3). The initiation of the second national lockdown was associated with an increase or no significant change in primary HCU (% increase (95%CI): new prescriptions: -0.3% (−1.3%-0.6%); BP: 4.1% (2.2%-6.1%); BMI: 2.4% (0.0%-4.8%); Cholesterol: 10.0% (7.3%-12.8%); HbA1c: 6.5% (4.1%-8.9%); Smoking status: -0.2% (−2.1%-1.8%)).

**Figure 1:**
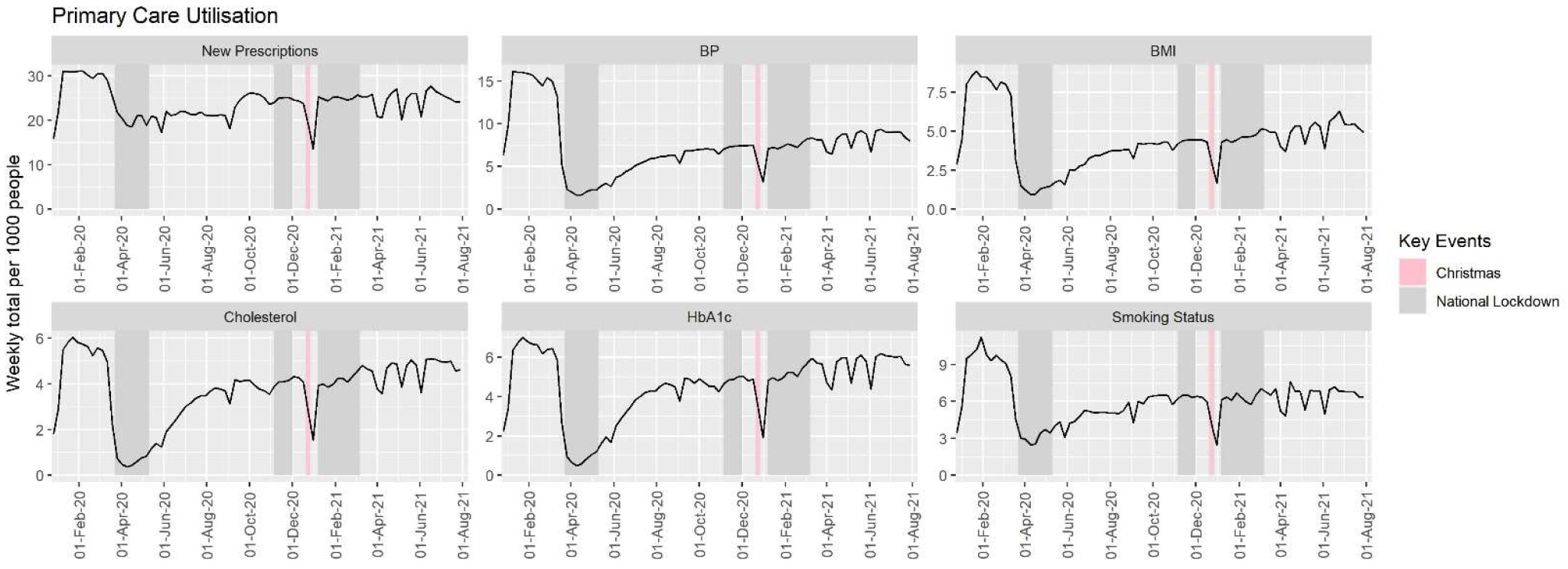
Weekly primary care utilisation per 1000 people of the Greater Manchester population between January 2020 and August 2021. The first week covered 30^th^ December 2019 to 5^th^ January 2020, hence utilisation was expected to be considerably lower between this and the following week due to the UK bank holiday and seasonal effects expected for this calendar week.

All primary care HCU moderately increased weekly from the initiation of the first national lockdown to the end of the study (RR (95%CI): new prescriptions: 1.00 (1.00 - 1.00; p<0.001); BP: 1.01 (1.01 – 1.01;p<0.001); BMI: 1.01 (1.01 – 1.01;p<0.001); Cholesterol: 1.02 (1.02 – 1.02;p<0.001); HbA1c: 1.01 (1.01 – 1.02;p<0.001); Smoking status: 1.01 (1.01 – 1.01;p<0.001)). Despite this, by the end of the study, all measures were still recorded less often than in the pre-pandemic period (RR (95% CI; p-value); new prescriptions: 0.828 (0.825 – 0.832; p<0.001); BP: 0.583 (0.580-0.587; p<0.001); BMI: 0.669 (0.664-0.675; p<0.001); cholesterol: 0.896 (0.888-0.905; p<0.001); HbA1c: 0.935 (0.927-0.943; p<0.001); smoking status: 0.711 (0.706-0.717; p<0.001)).

All primary care measures were lower across calendar weeks 2-11 when comparing 2021 data with 2020 data (p<0.001; Supplementary Table S4). The measuring of BP and BMI remained consistently lower throughout these weeks by an average of 50.0% (95%CI: 49.8%-50.2%; p<0.001) and 42.5% (95% CI: 42.1%-42.8%; p<0.001), respectively. Even though the rates of cholesterol and HbA1c measurements taken in calendar week 11 were similar in 2021 (pandemic) and 2020 (pre-pandemic): 0.937 (95% CI: 0.916 – 0.958) and 0.973 (95% CI: 0.953 – 0.992), respectively, they were still significantly lower in 2021.

### Primary HCU by multi-morbidity and deprivation

Multi-morbid patients and patients with one LTC had consistently higher levels of primary HCU than patients with no LTCs throughout the study period (Supplementary Table 3a; Figure 2). The ratio of weekly HCU rates per 1000 people between multi-morbid patients and those with no LTCs significantly increased for new prescriptions (RR: 1.281; 95%CI: 1.169 – 1.404; p <0.001), BP (RR: 1.187; 95%CI: 1.007 – 1.400; p = 0.042) and smoking status (RR: 1.356; 95%CI: 1.126 – 1.632; p = 0.002) during the first national lockdown, and decreased for BMI (RR: 0.736; 95%CI 0.613 – 0.885; p=0.001) and smoking status (RR: 0.803; 95%CI 0.675 – 0.956; p=0.014) in the third national lockdown. No significant changes in HCU between multi-morbid patients and those with no LTCs were observed for HBA1c or cholesterol or during the second national lockdown for all primary care HCU (Supplementary Table S6; Supplementary Figure S1).

**Figure 2:**
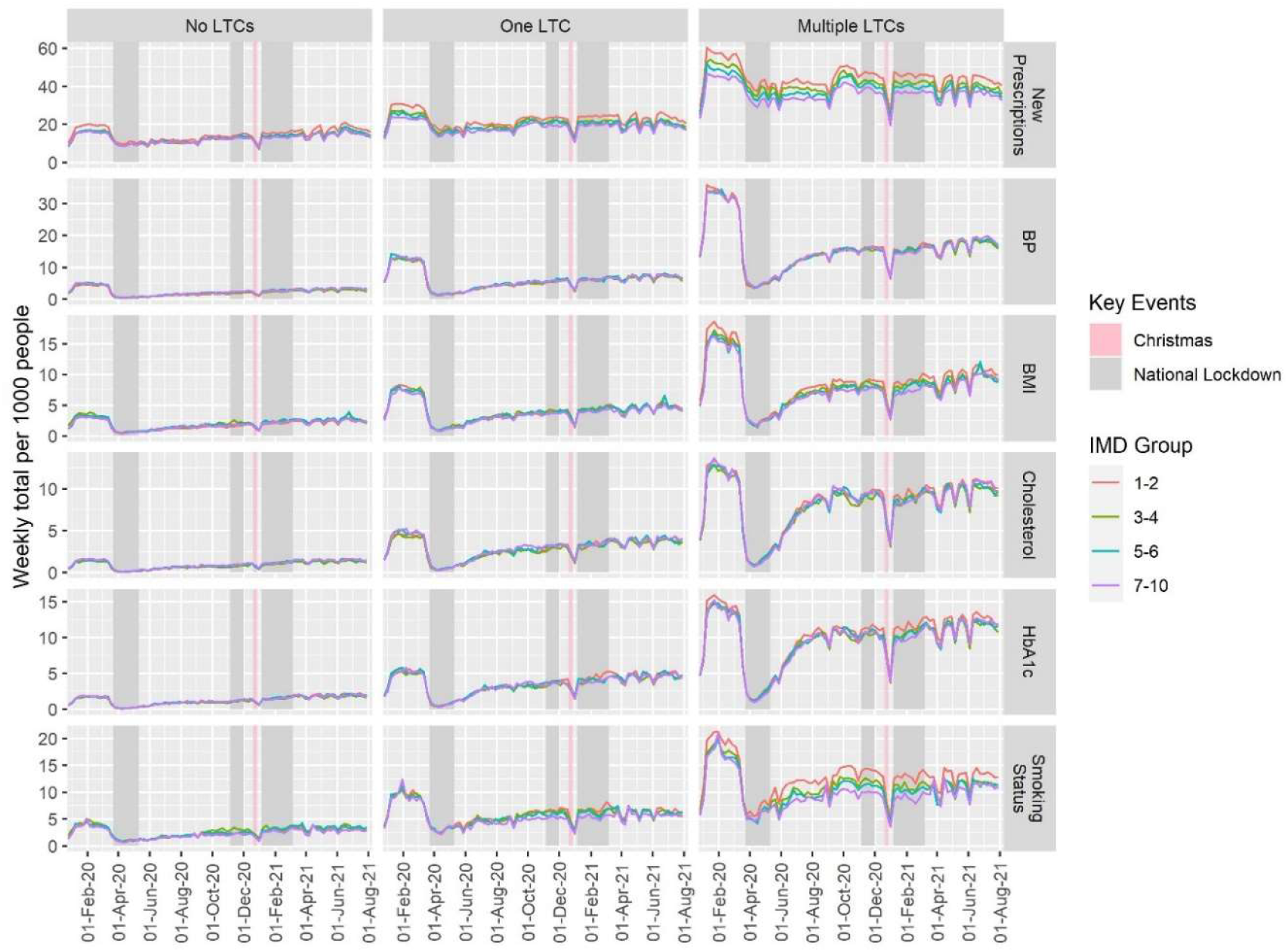
Rates of primary care measures recorded per 1000 people per week, identified according to number of long-term conditions and deprivation group, between January 2020 and August 2021.

### Primary HCU by deprivation

People that were less deprived had lower rates of new prescriptions compared to the most deprived group (IMD 1-2), (RR (95%CI); 3-4 vs 1-2: 0.915 (0.874 – 0.959); 5-6 vs 1-2: 0.920 (0.878 – 0.964); 7-10 vs 1-2: 0.875 (0.835 – 0.917); Figure 2, Supplementary Table S7). Similarly, smoking status had a lower rate of measurement in the least deprived patients compared to the most deprived patients (RR: 0.885; 95% CI: 0.801 – 0.877). No other differences were observed with regards to deprivation across primary HCU. The least deprived group had experienced an additional reduction in smoking status during the third national lockdown (RR: 0.836; 95%CI 0.704 - 0.994; p=0.042) but no other interactions between deprivation and national lockdowns were evident (Supplementary Table S9).

### Interaction between multi-morbidity and deprivation for primary HCU

Differences in HCU by deprivation were overall larger within multi-morbid patients (Supplementary Table S8; Figure 2). Differences in HCU between deprivation groups were not attributable to only one LTC group (Supplementary Figure S2). In multi-morbid patients, there were no significant changes in the ratio of weekly HCU per 1000 people between the least deprived group and the other deprivation groups across all primary HCU measures during the first national lockdown compared to pre-pandemic weeks (Supplementary Table S10; Supplementary Figure S3).

### Overall secondary HCU

There has been large variation in planned and unplanned secondary HCU over the course of the COVID-19 pandemic (Supplementary Figure S4). There was a 47.4% (95%CI: 42.9% - 51.5%, p<0.001) reduction in planned and 35.3% (95%CI: 28.3% - 41.6%; p<0.001) reduction in unplanned weekly admission rates per 1000 people associated with the initiation of the first national lockdown (planned: 2.51 to 1.32; unplanned: 1.36 to 0.88). The initiation of the second national lockdown was also associated with a significant reduction in secondary HCU; planned weekly admission rates per 1000 people reduced by 20.4% (95%CI 14.4%-25.9%; p<0.001) and unplanned reduced by 15.6% (95%CI 7.3%-23.1%; p<0.001). The reductions were sustained throughout these lockdowns; only unplanned admissions in the third national lockdown were not significantly lower than in pre-pandemic rates (Supplementary Table S3). The patterns observed in secondary admissions were consistent across all three main contributing secondary care providers (Supplementary Figure S5).

Both planned and unplanned weekly admissions were on average lower from the beginning of the first national lockdown up until the end of the study period, compared to the pre-pandemic admissions (planned: RR 0.850, 95%CI 0.837 – 0.864, p<0.001; unplanned: RR 0.976, 95%CI 0.957 – 0.996, p=0.016). However, the admissions increased throughout the period (planned: p<0.001; unplanned: p<0.001) and when comparing the final four weeks of the study period with the pre-pandemic period, planned admission rates were not significantly different (RR: 1.105; 95% CI 0.987 – 1.044; p=0.290) and unplanned were higher (RR: 1.104, 95% CI 1.067 – 1.143; p<0.001). The direct comparison between calendar weeks 2-11 in 2021 vs 2020, indicated that planned admissions were lower on average by 11.3% (95% CI: 9.4% - 13.2%; p<0.001) but there was no difference in unplanned admissions (RR: 1.012; 95%CI 0.985 – 1.040; p=0.376). A week-by-week comparison is detailed in Supplementary Table S3.

### Secondary HCU by multi-morbidity

Morbidity was associated with an increased rate of planned admissions throughout the study period: One vs No LTCs RR 1.904 (95% CI: 1.717 – 2.111; p<0.001); Multiple vs No LTCs RR 9.584 (95%CI 8.644 – 10.627; p<0.001); Multiple vs One LTC RR 5.033 (95%CI 4.540 – 5.581; p<0.001). This was also the case for unplanned admissions: One vs No LTCs RR 1.188 (95% CI: 1.112 – 1.270; p<0.001); Multiple vs No LTCs RR 3.636 (95%CI 3.401 – 3.887; p<0.001); Multiple vs One LTC RR 3.059 (95%CI 2.862 – 3.271; p<0.001) (Figure 3).

**Figure 3:**
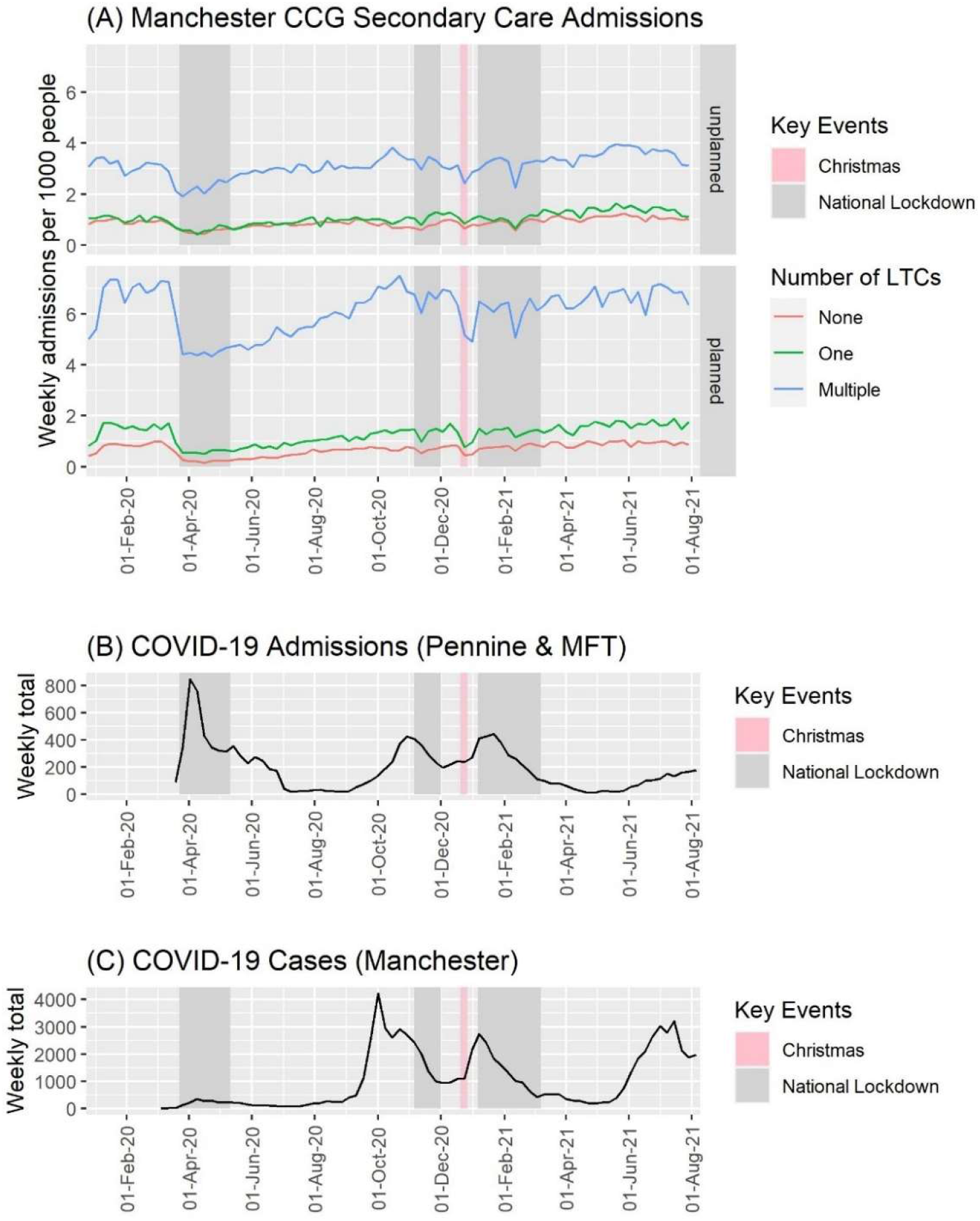
(A:) Weekly rates of planned and unplanned admissions per 1000 people that were identified as having zero, one or multiple LTCs, within the Manchester CCG subpopulation between January 2020 and August 2021. (B:) Government reported COVID-19 admissions in Manchester University NHS Foundation Trust, Pennine Acute Hospitals NHS Foundation Trust and Pennine Care NHS Foundation Trust (extracted 08-Sept-21), and (C:) Government reported cases in Manchester (extracted 08-Sept-21). *MFT = Manchester University Hospital Foundation Trust, Pennine = Pennine Acute Hospitals NHS Trust & Pennine Care NHS Foundation Trust*

Whilst the ratio of weekly unplanned admissions per 1000 people between patients that were multi-morbid vs those without any LTCs was consistent throughout the majority of the pandemic, there was a significant increase during the first lockdown compared with pre-pandemic (RR 1.253, 95%CI 1.068 – 1.469; p=0.006) but no significant change was observed between those with one LTC and those without any LTCs (RR 0.987, 95%CI 0.842 – 1.157; p=0.865, Supplementary Figure S6). The ratio of planned admission rates per 1000 people in morbidity groups increased during the first national lockdown compared to that observed pre-pandemic: multi-morbid vs no LTC increased by a factor of 2.402 (95% CI 2.047 – 2.818; p<0.001) and one LTC vs no LTCs increased by a factor of 1.413 (95% CI 1.205 – 1.658; p<0.001) (Supplementary Table S5; Supplementary Figure S6), however this was not sustained throughout the pandemic. The average of the ratios of admission rates between multi-morbid patients vs patients without LTCs from the start of the first national lockdown until the end of the study period vs pre-pandemic was 1.176 (95% CI: 0.871 – 1.588; p =0.289) for planned admission rates and 1.097 (95%CI: 0.900 – 1.338; p=0.357) for unplanned admission rates.

### Secondary care HCU for specific LTC groups

There were noticeable differences for both planned and unplanned admission rates within each LTC group over the study period (Figure 4). Planned admission rates were highest for patients with a renal or urological LTC. Unplanned admission rates were highest in patients with cancer or renal LTCs. Planned and unplanned admission rates were lowest overall for patients without any LTC. For cancer patients, the drop in the number of planned admissions at the initiation of the first national lockdown was sustained throughout the remainder of the study period, with an average reduction of 28.9% (95% CI: 25.1% - 32.5%; p<0.001) compared to pre-pandemic levels and unplanned admission rates decreased by 11.5% (95% CI 2.4% - 19.6%; p=0.014). Planned admission rates for people identified as having an endocrine, musculoskeletal or skin, neurological, psychiatric, or respiratory LTC returned to pre-pandemic levels by the end of the study period. However, planned admission rates for people identified as having cancer, cardiovascular, gastrointestinal, renal or urological and sensory impairment or learning disability remained lower than in the pre-pandemic period. Conversely, planned admission rates for people identified with a substance abuse LTC were higher by the end of the study period compared to the pre-pandemic period (RR: 1.196; 95%CI 1.07 – 1.329; p=0.001; Supplementary Table S11). Unplanned admissions rates were lower only for those that were identified with a renal or urological LTC. Patient groups with a gastrointestinal, musculoskeletal or skin, psychiatric or substance abuse LTC had higher rates of unplanned admissions at the end of the study compared to pre-pandemic levels. The remaining LTC groups had no significant change in unplanned admissions (Supplementary Table S11).

**Figure 4:**
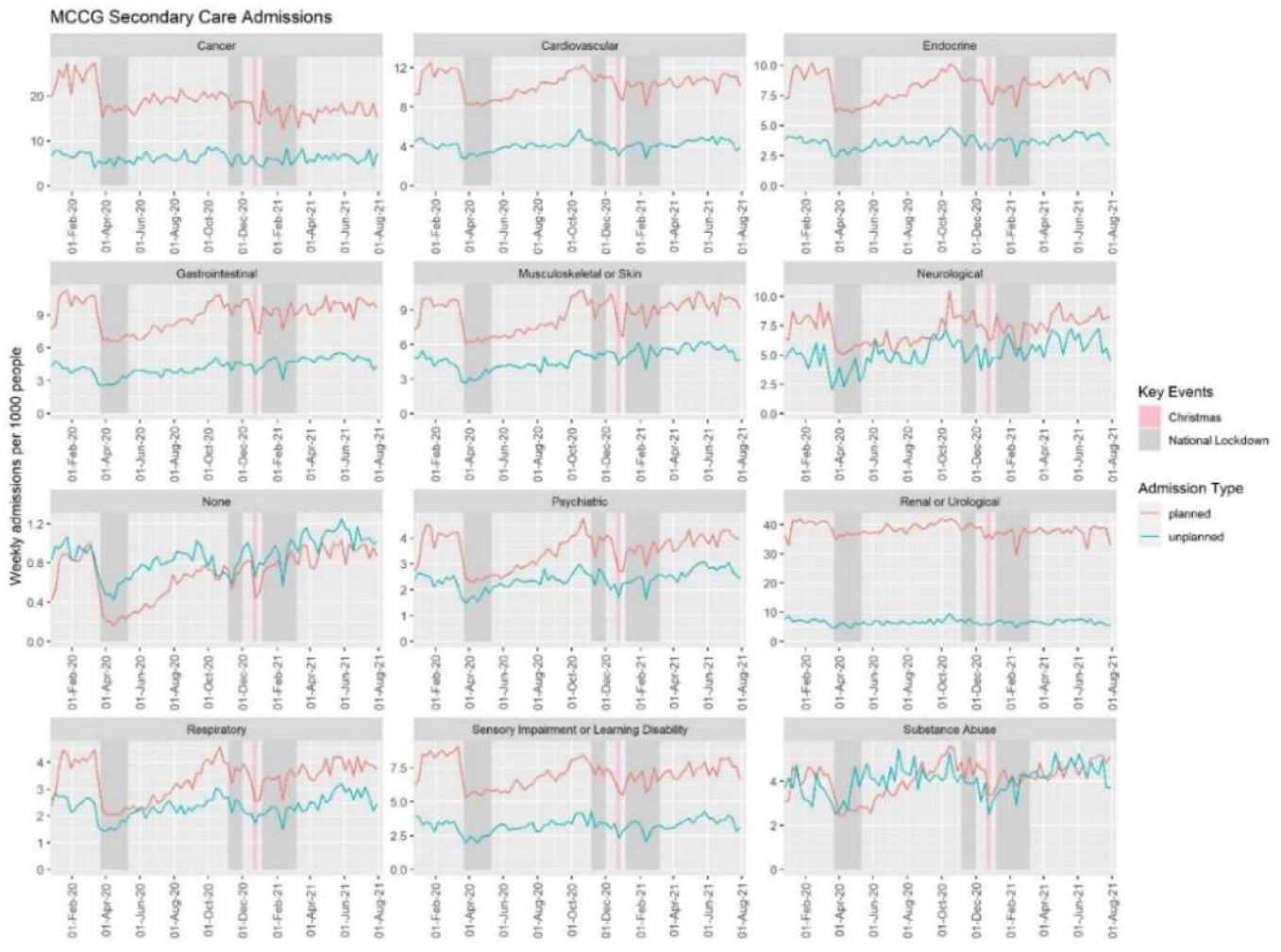
Weekly rates of planned and unplanned admissions identified in patients with each of the long-term conditions within the Manchester CCG subpopulation, between January 2020 and August 2021.

### Secondary HCU by deprivation

People that were most deprived (IMD of 1 or 2) had the highest rates for both planned (RR (95%CI); 3-4 vs 1-2: 0.753 (0.694 – 0.818; p<0.001); 5-6 vs 1-2: 0.787 (0.724 – 0.854;p<0.001); 7-10 vs 1-2: 0.812 (0.748 – 0.882; p<0.001)) and unplanned admissions (RR (95%CI); 3-4 vs 1-2: 0.686 (0.642 – 0.732; p<0.001); 5-6 vs 1-2: 0.670 (0.628 – 0.715; p<0.001); 7-10 vs 1-2: 0.683 (0.640 – 0.729; p<0.001)) throughout the study period. For multi-morbid patients, being highly deprived was associated with an increased rate in both planned and unplanned admissions compared to all other deprivation groups (Supplementary Table S3c; Supplementary Figure S7). The ratios of rates between deprivation groups within multi-morbid patients were not significantly different during the first national lockdown compared to pre-pandemic levels (Supplementary Table S10, Supplementary Figure S8).

## DISCUSSION

### Principle Findings and Interpretation

We have assessed primary HCU for over 3 million patients across GM and secondary HCU for a sub-group of almost 700,000 patients within Manchester CCG. Major changes in HCU occurred during the COVID-19 pandemic. There was a large reduction in both primary and secondary HCU at the beginning of the first national lockdown. Whilst there was a relatively consistent increase in primary care HCU from the first national lockdown, primary HCU remained was lower at the end of the study compared to pre-pandemic. Overall, both planned and unplanned secondary admissions had recovered to pre-pandemic levels by the end of the study period but this recovery was not observed across all LTC subgroups. Changes in the ratio of HCU between multi-morbid patients and those without LTCs occurred during national lockdowns but were inconsistent across primary HCU measures.

Although some healthcare information measures can be completed remotely (e.g. smoking status, BP and BMI), primary HCU measures that require in-person contact with a healthcare professional (e.g. HbA1c and cholesterol) demonstrated similar patterns in HCU. The initial larger fall in healthcare information recording compared to incident prescribing in primary care may reflect a shift in focus away from secondary prevention during the first wave of the pandemic. Although these HCU measures have not returned to pre-pandemic levels, they have consistently increased since the first lockdown and this has occurred even though quality outcome framework targets and local enhanced services were largely suspended.

Despite a peak in COVID-19 admissions within the first national lockdown, secondary admissions fell by a larger volume. Reductions in secondary care admissions associated with the first lockdown have been reported across the UK.^11,24,25^ Largely, these are reflective of cancellations of elective activity or delaying non-urgent care, to ensure capacity for patients with severe COVID-19 infection and to increase critical care capacity.^7^ The observed deficit may not correspond entirely to an unmet need of patients with non-COVID-19 healthcare needs as there is some evidence that changes in behaviour according to sanitisation campaigns, social distancing and government restrictions may have resulted in fewer infections,^26^ and injuries.^27^ Additionally, emergency department attendances which are related to unplanned admissions (but were not directly assessed in this study) have been observed to have reduced.^14^ It is also possible that the increased utilisation of remote management for secondary care patients has contributed to clinically appropriate reductions in admissions.

There has been no noticeable recovery in HCU for patients with cancer and for a number of other LTCs, recovery to pre-pandemic HCU levels has not occurred. In contrast, HCU of patients identified with substance abuse and/or a psychiatric condition exceeded pre-pandemic levels between the first and second national lockdowns, likely reaffirming the significant impact of the pandemic on mental health and psychiatric services.^28^

### Implications for Clinicians and Policy Makers

It is inevitable that the changes in HCU observed in this study will have had an impact on both patients and healthcare providers above and beyond the direct impact of COVID-19. For patients with cancer, services had to adapt to mitigate the increased risk of death from COVID-19.^29^ The initial reduction in the number of planned admissions was sustained throughout the study period and is likely to reflect changes in services but may also be due to patients with cancer being reluctant to seek healthcare. Delays in care for patients with cancer are known to impact prognosis,^30^ and the pandemic has been found to have contributed to excess deaths in patients with cancer.^24^ A proactive approach to encourage patients to attend screening and routine appointments will be needed to minimise the impact of the pandemic on patients with cancer and other emerging health inequalities.^31,32^ Understanding the implications of reductions in the selected primary care HCU measures, particularly the decrease in assessing and recording healthcare information will require further long-term studies.

### Strengths & Limitations

The strengths of this study include the complete coverage of a large geographical area for the primary care analyses and the inclusion of both primary and secondary HCU data. This is the first study to evaluate HCU across the full spectrum of LTC subpopulations and stratify according to multi-morbidity and deprivation. Data prior to 2020 were not available and consequently comparisons made (pre- vs post-pandemic) are reliant on the data between January and March 2020 being representative of pre-pandemic utilisation. Consequently, the comparison of pre-pandemic HCU to the end of the study period may have been influenced by seasonal variations in HCU. The secondary care analysis was only possible on a subset of the GM population due to delays in data from some GM secondary care providers. The study population represents a highly deprived population placed under strict restrictions during the pandemic. While this information is valuable, the findings may not be generalizable to other settings in the UK or internationally.

Although the measures of HCU that have been selected are relevant and reliable, they do not provide a complete picture of either primary or secondary HCU. There is no single effective measure to summarise HCU in primary care as there are many aspects that reflect HCU in this setting.^35^ It remains possible that the shift towards increased remote consultations may have resulted in changes to primary care delivery that were not possible to accurately capture using our measures of primary HCU. Additionally, the cause of admission was not available for secondary HCU, hence we were unable to determine LTC-specific admissions. Whilst the current scaling and sub-populations do not take into consideration any deaths or new diagnoses that occurred after 1 Jan 2020, a sensitivity analysis accounting for deaths resulted in very small increases to rates (Supplementary Figures S9-S10; Supplementary Table S12) and scaled utilisation remained lower than pre-lockdown levels.

## Conclusions

We have assessed the changes in HCU in primary and secondary care associated with the COVID-19 pandemic and UK national lockdowns for patients with LTCs across a large urban region. There was a significant reduction in both primary and secondary HCU associated with the first national lockdown. Subsequent national lockdowns were associated with reductions in secondary care but not in primary care. Whilst some measures of healthcare utilisation had returned to pre-pandemic levels by the end of the study, many had not. Proportionally, secondary care HCU increased in multi-morbid patients compared to those without LTCs during the first and second national lockdowns. Although changes to HCU during the pandemic have been similar overall, different patterns have been seen in specific LTC groups such as people with cancer. Over the course of the pandemic deprivation was associated with higher rates of HCU in multi-morbid patients but no significant differences were observed in the ratio of utilisation between the most and least deprived groups for the majority of HCU measures during national lockdowns.

## Data Availability

Data from the Greater Manchester Care Record is not freely available. Access to the data would need approval from the data provider. Requests to access the data should be directed to

## ACKNOWLEDGEMENTS

The time of RW and NP was partially funded by the National Institute for Health Research (NIHR) Greater Manchester Patient Safety Translational Research Centre (PSTRC-2016-003) and the NIHR Applied Research Collaboration Greater Manchester (NIHR200174). The time of NP was partially funded by the NIHR Manchester Biomedical Research Centre (IS-BRC-1215-20007). The time of CSP was fully funded by the NIHR Applied Research Collaboration Greater Manchester (NIHR200174). The views expressed are those of the author(s) and not necessarily those of the NIHR or the Department of Health and Social Care. The authors recognise the Greater Manchester Care Record (a partnership of Greater Manchester Health and Social Care Partnership, Health Innovation Manchester and Graphnet Health, on behalf of Greater Manchester localities) in the provision of data required to undertake this work. Using patient data is vital to improve health and care for everyone. There is huge potential to make better use of information from people’s patient records, to understand more about disease, develop new treatments, monitor safety, and plan NHS services. Patient data should be kept safe and secure, to protect everyone’s privacy, and it’s important that there are safeguards to make sure that it is stored and used responsibly. Everyone should be able to find out about how patient data is used.

## CONTRIBUTIONS

SG conceived the study. NP and RW led on the acquisition of data for the study. All authors were involved in the design of the study and interpretation of the data. CSP led the analysis of the data, supported by MS. CSP and SG prepared the manuscript. All authors revised the manuscript and approved the final version.

## COMPETING INTERESTS

All authors have completed the ICMJE uniform disclosure form at *http://www.icmje.org/disclosure-of-interest/* and declare: no support from any organisation for the submitted work; no financial relationships with any organisations that might have an interest in the submitted work in the previous three years; no other relationships or activities that could appear to have influenced the submitted work.

## Notes

### Competing Interest Statement

The authors have declared no competing interest.

### Author Declarations

The University of Manchester is permitted to perform research on this data via a Greater Manchester wide data protection impact assessment (DPIA). The basis for this DPIA is the control of patient information (COPI) notice issued by the Secretary of State for Health and Social Care in March 2020 which allowed confidential patient information to be shared for the purposes of research into COVID-19.17 The study was approved by both the GMCR Expert Review Group and Research Governance Group. All data made available to the analysts was de-identified and aggregated and therefore did not require specific ethical approval.

