## Supplementary Materials for "HEALTHCARE UTILISATION IN PATIENTS WITH LONG-TERM CONDITIONS DURING THE COVID-19 PANDEMIC: A POPULATION BASED STUDY ACROSS GREATER MANCHESTER, UK"

**Supplementary Table S1:** Long-term medical conditions and their groupings

| Grouping | Long-term medical condition |
|---|---|
| **Cardiovascular** | Hypertension<br>Atrial fibrillation<br>Heart failure<br>Peripheral vascular disease<br>Stroke & transient ischaemic attack<br>Coronary heart disease |
| **Respiratory** | Bronchiectasis<br>Asthma<br>Chronic obstructive pulmonary disease<br>Chronic sinusitis |
| **Gastrointestinal** | Viral Hepatitis<br>Chronic liver disease<br>Inflammatory bowel disease<br>Diverticular disease of intestine<br>Treated constipation<br>Irritable bowel syndrome<br>Treated dyspepsia |
| **Neurological** | Multiple sclerosis<br>Parkinson's disease<br>Migraine<br>Epilepsy |
| **Endocrine** | Thyroid disorders<br>Diabetes |
| **Psychiatric** | Anorexia or bulimia<br>Schizophrenia (and related non-organic psychosis) or bipolar disorder<br>Dementia<br>Anxiety & other neurotic, stress related & somatoform disorders<br>Depression |
| **Substance Abuse** | Alcohol problems<br>Other psychoactive substance misuse |
| **Musculoskeletal/Skin** | Psoriasis or eczema<br>Rheumatoid arthritis, other inflammatory polyarthropathies & systematic connective tissue disorders<br>Painful condition |
| **Sensory impairment or learning disability** | Learning disability<br>Blindness & low vision<br>Glaucoma<br>Hearing loss |
| **Renal/Urological** | Chronic kidney disease<br>Prostate disorders |

**COVID-19 Government data extraction**

The Government data were extracted on 08-09-2021 from https://api.coronavirus.data.gov.uk/ . For the new cases, we extracted data for Manchester only. The API for Manchester cases by specimen date that was used is:

https://api.coronavirus.data.gov.uk/v2/data?areaType=utla&areaCode=E08000003&metric=newCasesBySpecimenDate&format=csv

For the secondary admissions, we exported the data for the trusts which were the main providers in the Manchester CCG subpopulation. The hospital groupings were not consistent across data sources; Table S2 provides the mapping between the Government listed NHS trusts and the secondary providers covering nearly all admissions experienced by Manchester CCG population in the GMCR.

**Supplementary Table S2**: Mapping between NHS acute providers according to data source

| GMCR acute provider | Government listed NHS Trust | Government data API |
|---|---|---|
| University Hospital of South Manchester | Manchester University NHS Foundation Trust | https://api.coronavirus.data.gov.uk/v2/data?areaType=nhsTrust&areaCode=R0A&metric=newAdmissions&format=csv |
| Central Manchester University Hospitals | | |
| Pennine Acute Hospitals | Pennine Acute Hospitals NHS Trust | https://api.coronavirus.data.gov.uk/v2/data?areaType=nhsTrust&areaCode=RW6&metric=newAdmissions&format=csv |
| Pennine Acute Hospitals | Pennine Care NHS Foundation Trust | https://api.coronavirus.data.gov.uk/v2/data?areaType=nhsTrust&areaCode=RT2&metric=newAdmissions&format=csv |

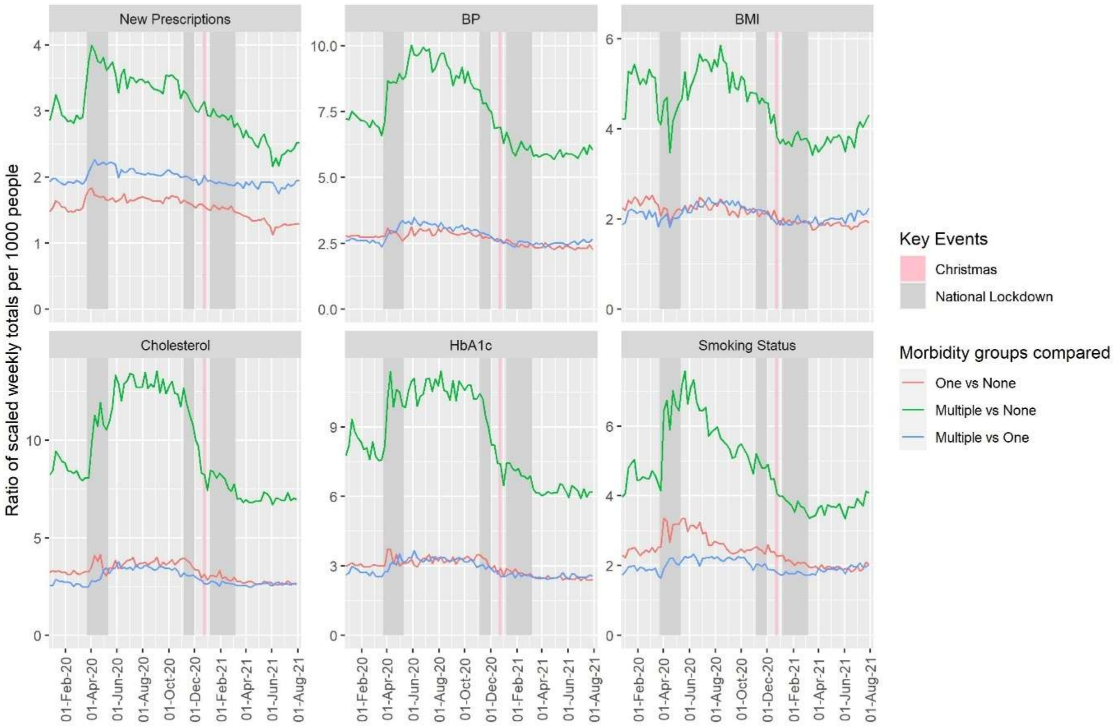


**Supplementary Figure S1:** Ratio of weekly primary care HCU measures per 1000 people between morbidity groups, between January 2020 and August 2021.

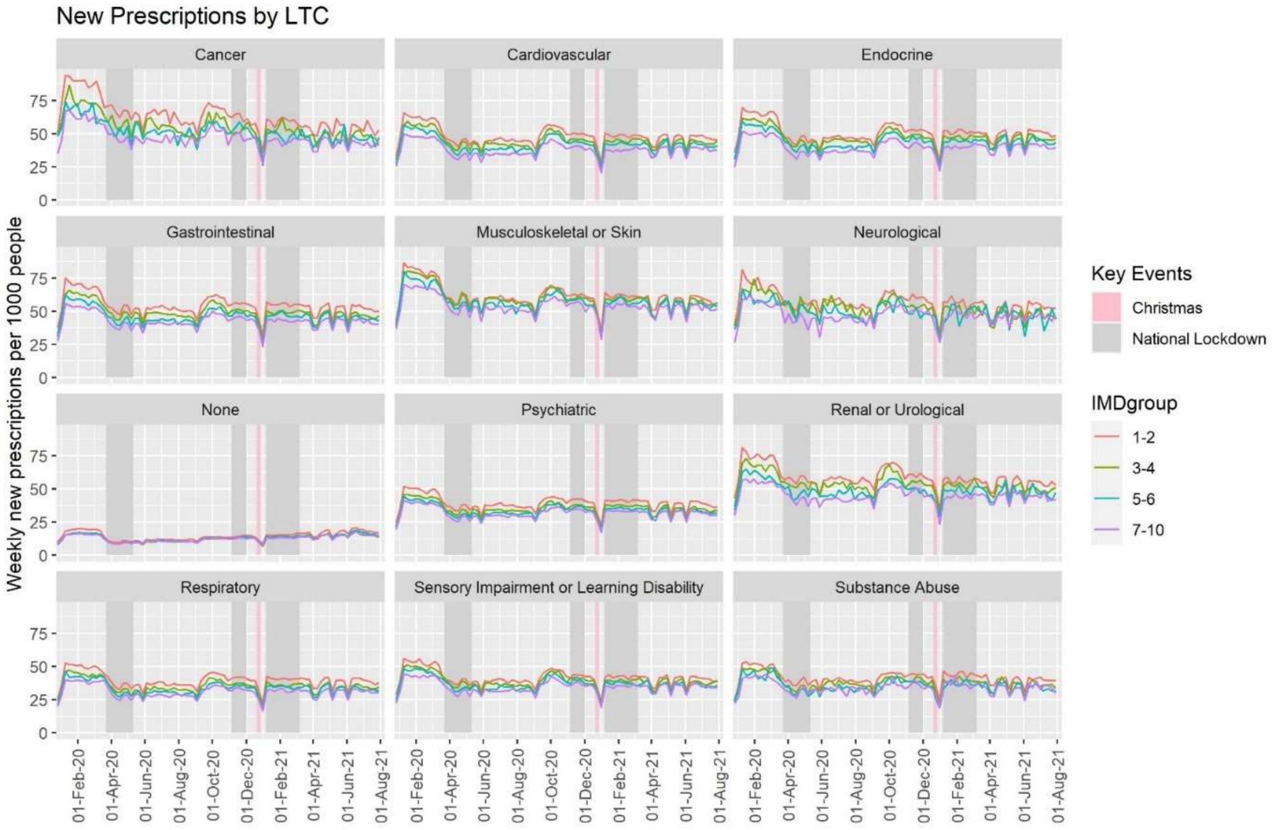


**Supplementary Figure S2:** Weekly new primary care prescriptions per 1000 people within each long-term condition group and deprivation group, between January 2020 and August 2021.

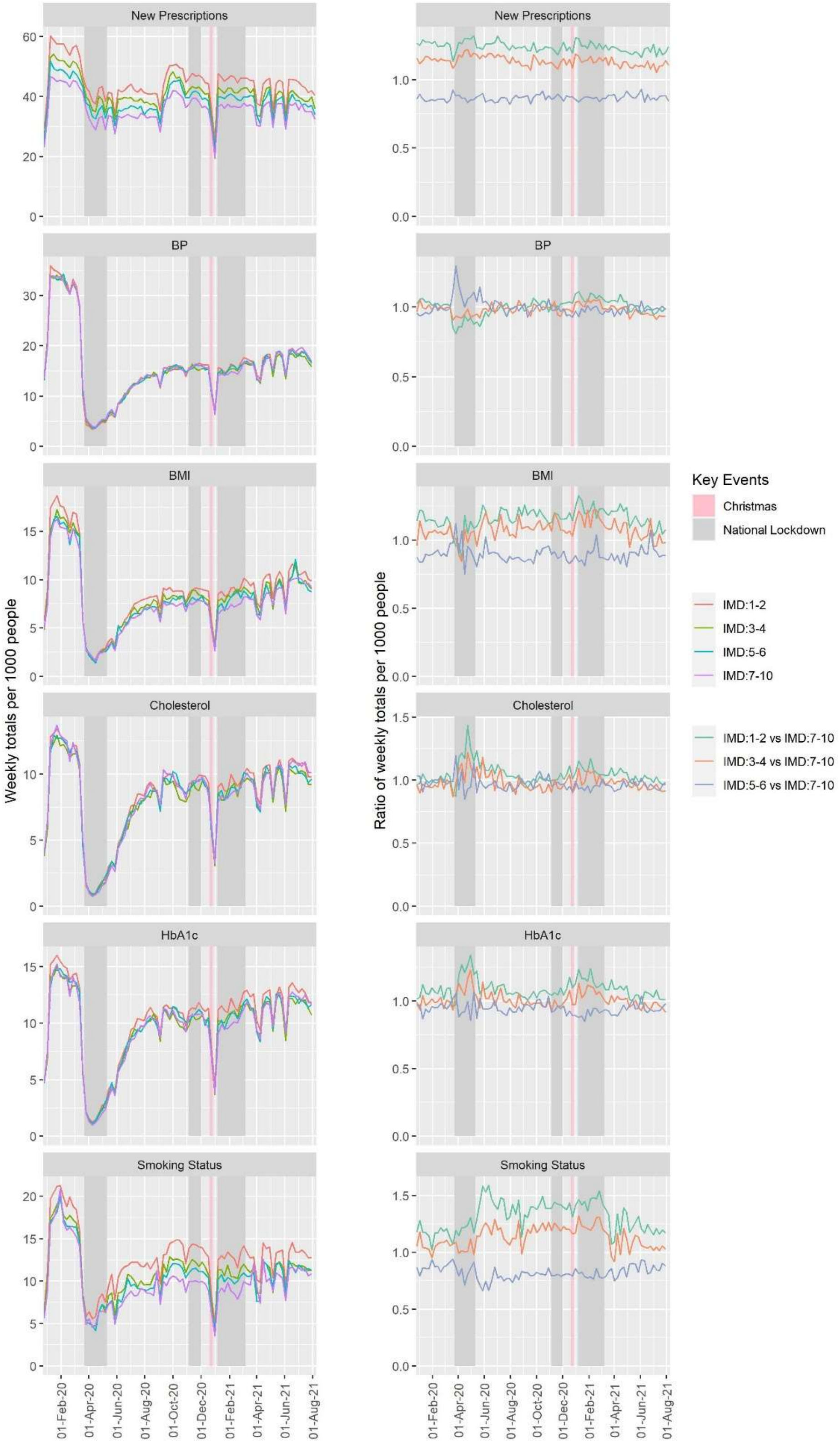


**Supplementary Figure S3:** Weekly new primary care HCU measures per 1000 people within multi-morbid patients by deprivation group and the ratio of these compared to the least-deprived group (IMD: 7-10), between January 2020 and August 2021.

**Supplementary Table S3:** Associated effects of the national lockdowns on primary HCU compare to pre-pandemic HCU. NL1 = First national lockdown, NL2 = Second national lockdown, NL3 = Third national lockdown.

| | Rate Ratio | 95% CI | p-value |
|---|---|---|---|
| | **New Prescriptions** | | |
| NL1 vs pre-pandemic | 0.681 | 0.679 – 0.684 | **<0.001** |
| NL2 vs pre-pandemic | 0.835 | 0.832 – 0.838 | **<0.001** |
| NL3 vs pre-pandemic | 0.843 | 0.841 – 0.846 | **<0.001** |
| | **BP** | | |
| NL1 vs pre-pandemic | 0.143 | 0.142 – 0.144 | **<0.001** |
| NL2 vs pre-pandemic | 0.494 | 0.491 – 0.497 | **<0.001** |
| NL3 vs pre-pandemic | 0.510 | 0.508 – 0.513 | **<0.001** |
| | **BMI** | | |
| NL1 vs pre-pandemic | 0.168 | 0.167 – 0.170 | **<0.001** |
| NL2 vs pre-pandemic | 0.559 | 0.555 – 0.564 | **<0.001** |
| NL3 vs pre-pandemic | 0.592 | 0.588 – 0.596 | **<0.001** |
| | **Cholesterol** | | |
| NL1 vs pre-pandemic | 0.128 | 0.126 – 0.130 | **<0.001** |
| NL2 vs pre-pandemic | 0.769 | 0.762 – 0.776 | **<0.001** |
| NL3 vs pre-pandemic | 0.788 | 0.782 – 0.793 | **<0.001** |
| | **HbA1c** | | |
| NL1 vs pre-pandemic | 0.148 | 0.146 – 0.150 | **<0.001** |
| NL2 vs pre-pandemic | 0.785 | 0.778 – 0.791 | **<0.001** |
| NL3 vs pre-pandemic | 0.837 | 0.832 – 0.842 | **<0.001** |
| | **Smoking Status** | | |
| NL1 vs pre-pandemic | 0.346 | 0.344 – 0.349 | **<0.001** |
| NL2 vs pre-pandemic | 0.692 | 0.687 – 0.696 | **<0.001** |
| NL3 vs pre-pandemic | 0.689 | 0.685 – 0.692 | **<0.001** |
| | **Planned Admissions** | | |
| NL1 vs pre-pandemic | 0.544 | 0.530 – 0.559 | **<0.001** |
| NL2 vs pre-pandemic | 0.921 | 0.897 – 0.946 | **<0.001** |
| NL3 vs pre-pandemic | 0.902 | 0.883 – 0.922 | **<0.001** |
| | **Unplanned Admissions** | | |
| NL1 vs pre-pandemic | 0.659 | 0.638 – 0.681 | **<0.001** |
| NL2 vs pre-pandemic | 0.953 | 0.922 – 0.986 | **0.005** |
| NL3 vs pre-pandemic | 0.984 | 0.958 – 1.010 | 0.227 |

**Supplementary Table S4:** Percentage decrease (95%CI) in HCU in 2021 compared with the same calendar week in 2020.

| Calendar Week | 2 | 3 | 4 | 5 | 6 | 7 | 8 | 9 | 10 | 11 | Average | p-value |
|---|---|---|---|---|---|---|---|---|---|---|---|---|
| **Primary HCU** | | | | | | | | | | | | |
| New prescriptions | 20.0 (19.3 – 20.8) | 21.2 (20.5 – 21.9) | 18.4 (17.7 – 19.2) | 18.7 (18.0 – 19.5) | 19.6 (18.9 – 20.3) | 18.7 (17.9 – 19.5) | 15.5 (14.8 – 16.3) | 15.5 (14.8 – 16.3) | 17.2 (16.5 – 18.0) | 12.8 (12.0 – 13.6) | 17.8 (17.6 – 18.1) | <0.001 |
| Blood Pressure | 55.2 (54.5 – 55.9) | 55.8 (55.1 – 56.5) | 54.4 (53.7 – 55.1) | 52.0 (51.3 – 52.7) | 52.4 (51.6 – 53.1) | 52.1 (51.4 – 52.9) | 45.9 (45.0 – 46.7) | 46.4 (45.7 – 47.2) | 44.3 (43.4 – 45.1) | 38.7 (37.8 – 39.7) | 50.0 (49.8 – 50.2) | <0.001 |
| BMI | 44.4 (43.3 – 45.5) | 49.7 (48.7 – 50.8) | 49.7 (48.7 – 50.8) | 45.3 (44.3 – 46.5) | 45.7 (44.6 – 46.8) | 43.2 (42.0 – 44.3) | 37.8 (36.6 – 39.1) | 36.7 (35.6 – 38.0) | 36.3 (35.1 – 37.6) | 32.6 (31.3 – 34.0) | 42.5 (42.1 – 42.8) | <0.001 |
| Cholesterol | 27.5 (25.8 – 29.1) | 34.0 (32.4 – 35.4) | 34.2 (32.7 – 35.6) | 26.9 (25.3 – 28.5) | 26.2 (24.5 – 27.7) | 27.4 (25.8 – 29.1) | 17.1 (15.3 – 19.0) | 18.0 (16.2 – 19.8) | 11.9 (10.0 – 13.8) | 6.3 (4.2 – 8.4) | 18.4 (17.8 – 18.9) | <0.001 |
| HbA1c | 22.0 (20.4 – 23.6) | 28.6 (27.2 – 30.1) | 29.4 (27.9 – 30.8) | 23.0 (21.5 – 24.6) | 21.7 (20.1 – 23.2) | 24.1 (22.6 – 25.7) | 12.4 (10.6 – 14.2) | 10.7 (9.0 – 12.5) | 7.7 (5.9 – 9.5) | 2.7 (0.8 – 4.7) | 18.6 (18.1 – 19.1) | <0.001 |
| Smoking Status | 33.3 (32.1 – 34.5) | 37.8 (36.7 – 38.9) | 34.7 (33.6 – 35.9) | 43.4 (42.6 – 44.5) | 38.9 (37.8 – 40.0) | 38.4 (37.3 – 39.6) | 32.9 (31.8 – 34.2) | 24.7 (23.5 – 26.1) | 25.1 (23.9 – 26.6) | 18.8 (17.4 – 20.4) | 33.3 (33.0 – 33.7) | <0.001 |
| **Secondary HCU** | | | | | | | | | | | | |
| Planned admissions | 11.6 (5.3 – 17.5) | 16.8 (10.9 – 22.3) | 12.5 (6.3 – 18.2) | -0.4 (-7.6 – 6.3) | 27.1 (21.6 – 32.3) | 12.5 (6.2 – 18.4) | 1.7 (-5.2 – 8.2) | 12.5 (6.3 – 18.3) | 14.8 (8.9 – 20.4) | 2.6 (-4.1 – 8.9) | 11.3 (9.4 – 13.2) | <0.001 |
| Unplanned admissions | 8.4 (0.2 – 15.9) | 3.3 (-5.2 – 11.2) | 11.2 (-7.4 – 9.1) | -14.2 (-24.9 – -4.4) | 26.9 (19.4 – 33.8) | 2.0 (-6.8 – 10.1) | -7.7 (-17.2 – 1.1) | -5.7 (-15.1 – 2.9) | -3.1 (-12.2 – 5.2) | -25.7 (-37.0 – -15.5) | -1.2 (-4.0 – 1.5) | 0.376 |

**Supplementary Table S5**: Associated effects of morbidity on the rates of weekly totals of primary and secondary HCU (per 1000 people), throughout the study period. LTC = Long term condition.

| | Rate Ratio | 95% CI | p-value |
|---|---|---|---|
| | **New Prescriptions** | | |
| Multiple LTCs vs No LTCs | 3.040 | 2.881 – 3.208 | <0.001 |
| One LTC vs No LTCs | 1.534 | 1.454 – 1.619 | <0.001 |
| | **BP** | | |
| Multiple LTCs vs No LTCs | 7.347 | 6.197 – 8.711 | <0.001 |
| One LTC vs No LTCs | 2.659 | 2.243 – 3.153 | <0.001 |
| | **BMI** | | |
| Multiple LTCs vs No LTCs | 4.429 | 3.801 – 5.162 | <0.001 |
| One LTC vs No LTCs | 2.106 | 1.807 – 2.454 | <0.001 |
| | **Cholesterol** | | |
| Multiple LTCs vs No LTCs | 9.360 | 7.654 – 11.446 | <0.001 |
| One LTC vs No LTCs | 3.226 | 2.638 – 3.945 | <0.001 |
| | **HbA1c** | | |
| Multiple LTCs vs No LTCs | 8.291 | 6.847 – 10.038 | <0.001 |
| One LTC vs No LTCs | 2.918 | 2.410 – 3.533 | <0.001 |
| | **Smoking Status** | | |
| Multiple LTCs vs No LTCs | 4.674 | 4.192 – 5.211 | <0.001 |
| One LTC vs No LTCs | 2.372 | 2.127 – 2.645 | <0.001 |
| | **Planned Admissions** | | |
| Multiple LTCs vs No LTCs | 9.584 | 8.644 – 10.627 | <0.001 |
| One LTC vs No LTCs | 1.904 | 1.717 – 2.111 | <0.001 |
| | **Unplanned Admissions** | | |
| Multiple LTCs vs No LTCs | 3.636 | 3.401 – 3.887 | <0.001 |
| One LTC vs No LTCs | 1.188 | 1.112 – 1.270 | <0.001 |

**Supplementary Table S6**: Estimated rate ratios (RRs) from log-linear regression models for each HCU measure observed pre-pandemic and in the national lockdowns, adjusted for the number of long term conditions (LTCs). NL1 = First national lockdown, NL2 = Second national lockdown, NL3 = Third national lockdown.

| | **Primary Care** | | | | | | | | | | | |
|---|---|---|---|---|---|---|---|---|---|---|---|---|
| | **New Prescriptions** | | | **BP** | | | **BMI** | | | **Cholesterol** | | |
| | RR | 95% CI | p-value | RR | 95% CI | p-value | RR | 95% CI | p-value | RR | 95% CI | p-value |
| NL1 vs pre-pandemic | 0.580 | 0.543 - 0.619 | **<0.001** | 0.126 | 0.112 - 0.141 | **<0.001** | 0.190 | 0.165 - 0.218 | **<0.001** | 0.103 | 0.084 - 0.127 | **<0.001** |
| NL2 vs pre-pandemic | 0.800 | 0.742 - 0.862 | **<0.001** | 0.469 | 0.409 - 0.537 | **<0.001** | 0.615 | 0.523 - 0.722 | **<0.001** | 0.624 | 0.490 - 0.796 | **<0.001** |
| NL3 vs pre-pandemic | 0.855 | 0.804 - 0.909 | **<0.001** | 0.581 | 0.520 - 0.648 | **<0.001** | 0.752 | 0.660 - 0.856 | **<0.001** | 0.859 | 0.705 - 1.046 | **0.128** |
| Multiple LTCs vs No LTCs | 2.950 | 2.779 - 3.131 | **<0.001** | 7.155 | 6.429 - 7.963 | **<0.001** | 5.091 | 4.483 - 5.781 | **<0.001** | 8.576 | 7.077 - 10.392 | **<0.001** |
| One LTC vs No LTCs | 1.531 | 1.442 - 1.625 | **<0.001** | 2.753 | 2.474 - 3.064 | **<0.001** | 2.400 | 2.114 - 2.726 | **<0.001** | 3.222 | 2.659 - 3.904 | **<0.001** |
| NL1 vs pre-pandemic : Multiple LTCs vs No LTCs | 1.281 | 1.169 - 1.404 | **<0.001** | 1.187 | 1.007 - 1.400 | **0.042** | 0.847 | 0.696 - 1.030 | 0.095 | 1.220 | 0.907 - 1.640 | 0.186 |
| NL2 vs pre-pandemic : Multiple LTCs vs No LTCs | 1.073 | 0.964 - 1.193 | 0.194 | 1.091 | 0.901 - 1.321 | 0.369 | 0.911 | 0.726 - 1.144 | 0.418 | 1.318 | 0.934 - 1.858 | 0.114 |
| NL3 vs pre-pandemic : Multiple LTCs vs No LTCs | 0.984 | 0.903 - 1.073 | 0.713 | 0.859 | 0.736 - 1.004 | 0.056 | 0.736 | 0.613 - 0.885 | **0.001** | 0.920 | 0.697 - 1.216 | 0.555 |
| NL1 vs pre-pandemic : One LTC vs No LTCs | 1.126 | 1.027 - 1.234 | **0.012** | 1.025 | 0.870 - 1.209 | 0.763 | 0.889 | 0.731 - 1.081 | 0.236 | 1.102 | 0.820 - 1.482 | 0.515 |
| NL2 vs pre-pandemic : One LTC vs No LTCs | 1.043 | 0.937 - 1.160 | 0.439 | 0.996 | 0.823 - 1.206 | 0.968 | 0.888 | 0.708 - 1.115 | 0.304 | 1.139 | 0.808 - 1.606 | 0.454 |
| NL3 vs pre-pandemic : One LTC vs No LTCs | 0.994 | 0.912 - 1.084 | 0.895 | 0.896 | 0.767 - 1.046 | 0.162 | 0.811 | 0.674 - 0.975 | **0.026** | 0.921 | 0.697 - 1.216 | 0.557 |
| | **Primary Care (ctd)** | | | | | | **Secondary Care** | | | | | |
| | **HbA1c** | | | **Smoking Status** | | | **Planned Admissions** | | | **Unplanned Admissions** | | |
| | RR | 95% CI | p-value | RR | 95% CI | p-value | RR | 95% CI | p-value | RR | 95% CI | p-value |

| | | | | | | | | | | | | |
|---|---|---|---|---|---|---|---|---|---|---|---|---|
| NL1 vs pre-pandemic | 0.120 | 0.096 - 0.148 | **<0.001** | 0.272 | 0.238 - 0.310 | **<0.001** | 0.272 | 0.243 - 0.305 | **<0.001** | 0.582 | 0.511 - 0.663 | **<0.001** |
| NL2 vs pre-pandemic | 0.716 | 0.557 - 0.920 | **0.010** | 0.667 | 0.573 - 0.777 | **<0.001** | 0.820 | 0.719 - 0.935 | **0.003** | 0.803 | 0.690 - 0.934 | **0.005** |
| NL3 vs pre-pandemic | 0.967 | 0.789 - 1.185 | 0.745 | 0.816 | 0.721 - 0.923 | **<0.001** | 0.935 | 0.841 - 1.040 | 0.214 | 0.933 | 0.826 - 1.055 | 0.267 |
| Multiple LTCs vs No LTCs | 8.223 | 6.744 - 10.026 | **<0.001** | 4.595 | 4.074 - 5.183 | **<0.001** | 8.298 | 7.481 - 9.205 | **<0.001** | 3.359 | 2.981 - 3.786 | **<0.001** |
| One LTC vs No LTCs | 3.022 | 2.478 - 3.684 | **<0.001** | 2.404 | 2.131 - 2.712 | **<0.001** | 1.834 | 1.653 - 2.034 | **<0.001** | 1.111 | 0.986 - 1.252 | 0.083 |
| NL1 vs pre-pandemic : Multiple LTCs vs No LTCs | 1.221 | 0.900 - 1.658 | 0.197 | 1.356 | 1.126 - 1.632 | **0.002** | 2.402 | 2.047 - 2.818 | **<0.001** | 1.253 | 1.042 - 1.506 | **0.017** |
| NL2 vs pre-pandemic : Multiple LTCs vs No LTCs | 1.145 | 0.803 - 1.633 | 0.449 | 1.074 | 0.866 - 1.332 | 0.511 | 1.174 | 0.975 - 1.413 | 0.090 | 1.283 | 1.036 - 1.588 | **0.023** |
| NL3 vs pre-pandemic : Multiple LTCs vs No LTCs | 0.852 | 0.639 - 1.136 | 0.271 | 0.803 | 0.675 - 0.956 | **0.014** | 0.957 | 0.824 - 1.112 | 0.563 | 1.073 | 0.902 - 1.276 | 0.421 |
| NL1 vs pre-pandemic : One LTC vs No LTCs | 1.082 | 0.797 - 1.468 | 0.610 | 1.279 | 1.062 - 1.540 | **0.010** | 1.413 | 1.205 - 1.658 | **<0.001** | 0.987 | 0.821 - 1.186 | 0.885 |
| NL2 vs pre-pandemic : One LTC vs No LTCs | 1.058 | 0.742 - 1.508 | 0.754 | 1.028 | 0.829 - 1.275 | 0.799 | 1.063 | 0.883 - 1.280 | 0.513 | 1.295 | 1.046 - 1.604 | **0.018** |
| NL3 vs pre-pandemic : One LTC vs No LTCs | 0.894 | 0.671 - 1.192 | 0.442 | 0.87 | 0.730 - 1.036 | 0.116 | 0.971 | 0.835 - 1.128 | 0.695 | 1.075 | 0.904 - 1.279 | 0.407 |

**Supplementary Table S7**: Associated effects of deprivation groups on primary and secondary HCU, throughout the study.

| | Primary Care | | | | | | | | | | | |
|---|---|---|---|---|---|---|---|---|---|---|---|---|
| Deprivation group compared to group 1-2 | **New Prescriptions** | | | **BP** | | | **BMI** | | | **Cholesterol** | | |
| | RR | 95% CI | p-value | RR | 95% CI | p-value | RR | 95% CI | p-value | RR | 95% CI | p-value |
| 3-4 | 0.915 | 0.874 – 0.959 | **<0.001** | 1.018 | 0.869 – 1.193 | 0.821 | 1.007 | 0.865 – 1.172 | 0.928 | 0.964 | 0.797 – 1.166 | 0.702 |
| 5-6 | 0.920 | 0.878 – 0.964 | **<0.001** | 1.117 | 0.954 – 1.309 | 0.169 | 1.025 | 0.881 – 1.192 | 0.751 | 1.076 | 0.889 – 1.302 | 0.450 |
| 7-10 | 0.875 | 0.835 – 0.917 | **<0.001** | 1.134 | 0.968 – 1.328 | 0.120 | 0.988 | 0.849 – 1.150 | 0.879 | 1.091 | 0.902 – 1.320 | 0.369 |
| | **Primary Care (ctd)** | | | | | | **Secondary Care** | | | | | |
| | **HbA1c** | | | **Smoking Status** | | | **Planned Admissions** | | | **Unplanned Admissions** | | |
| | RR | 95% CI | p-value | RR | 95% CI | p-value | RR | 95% CI | p-value | RR | 95% CI | p-value |
| 3-4 | 0.950 | 0.794 – 1.137 | 0.574 | 0.947 | 0.858 – 1.047 | 0.287 | 0.753 | 0.694 – 0.818 | <0.001 | 0.686 | 0.642 – 0.732 | <0.001 |
| 5-6 | 1.038 | 0.868 – 1.242 | 0.682 | 0.945 | 0.856 – 1.045 | 0.269 | 0.787 | 0.724 – 0.854 | <0.001 | 0.67 | 0.628 – 0.715 | <0.001 |
| 7-10 | 1.031 | 0.862 – 1.234 | 0.737 | 0.885 | 0.801 – 0.977 | 0.016 | 0.812 | 0.748 – 0.882 | <0.001 | 0.683 | 0.640 – 0.729 | <0.001 |

**Supplementary Table S8**: Comparison of HCU between deprivation groups and the highly deprived population throughout the pandemic for multi-morbid patients.

| | Deprivation 3-4 vs 1-2 | | Deprivation 5-6 vs 1-2 | | Deprivation 7-10 vs 1-2 | |
|---|---|---|---|---|---|---|
| | RR (95% CI) | p-value | RR (95% CI) | p-value | RR (95% CI) | p-value |
| **Primary HCU** | | | | | | |
| New prescriptions | 0.917 (0.878 – 0.957) | **<0.001** | 0.867 (0.830 – 0.905) | **<0.001** | 0.807 (0.773 – 0.843) | **<0.001** |
| BP | 0.983 (0.844 – 1.145) | 0.827 | 0.997 (0.856 – 1.161) | 0.969 | 1.001 (0.860 – 1.166) | 0.989 |
| BMI | 0.927 (0.796 – 1.081) | 0.334 | 0.893 (0.766 – 1.041) | 0.147 | 0.865 (0.742 – 1.008) | 0.063 |
| Cholesterol | 0.939 (0.779 – 1.133) | 0.510 | 0.969 (0.803 – 1.168) | 0.738 | 0.958 (0.794 – 1.155) | 0.652 |
| HbA1c | 0.927 (0.779 – 1.104) | 0.396 | 0.942 (0.791 – 1.122) | 0.504 | 0.919 (0.772 – 1.094) | 0.342 |
| Smoking status | 0.863 (0.785 – 0.949) | **0.003** | 0.820 (0.745 – 0.901) | **<0.001** | 0.766 (0.697 – 0.843) | **<0.001** |
| **Secondary HCU** | | | | | | |
| Planned admissions | 0.873 (0.773 – 0.986) | **0.029** | 0.741 (0.652 – 0.842) | **<0.001** | 0.705 (0.619 – 0.802) | **<0.001** |
| Unplanned admissions | 0.826 (0.696 – 0.980) | **0.029** | 0.669 (0.557 – 0.802) | **<0.001** | 0.645 (0.536 – 0.774) | **<0.001** |

**Supplementary Table S9**: Estimated rate ratios (RRs) from log-linear regression models for each HCU observed pre-pandemic and in the national lockdowns, adjusted for the deprivation group. NL1 = First national lockdown, NL2 = Second national lockdown, NL3 = Third national lockdown, IMD = index of multiple deprivation.

| | **Primary Care** | | | | | | | | | | | |
|---|---|---|---|---|---|---|---|---|---|---|---|---|
| | **New Prescriptions** | | | **BP** | | | **BMI** | | | **Cholesterol** | | |
| | RR | 95% CI | p-value | RR | 95% CI | p-value | RR | 95% CI | p-value | RR | 95% CI | p-value |
| NL1 vs pre-pandemic | 0.668 | 0.629 – 0.711 | **<0.001** | 0.133 | 0.118 – 0.149 | **<0.001** | 0.161 | 0.139 – 0.186 | **<0.001** | 0.125 | 0.102 – 0.154 | **<0.001** |
| NL2 vs pre-pandemic | 0.819 | 0.763 – 0.879 | **<0.001** | 0.491 | 0.430 – 0.560 | **<0.001** | 0.555 | 0.469 – 0.656 | **<0.001** | 0.782 | 0.615 – 0.994 | **0.045** |
| NL3 vs pre-pandemic | 0.837 | 0.790 – 0.886 | **<0.001** | 0.520 | 0.467 – 0.579 | **<0.001** | 0.599 | 0.523 – 0.686 | **<0.001** | 0.824 | 0.678 – 1.001 | 0.051 |
| IMD: 3-4 vs IMD: 1-2 | 0.895 | 0.846 – 0.947 | **<0.001** | 1.011 | 0.910 – 1.123 | 0.837 | 0.994 | 0.871 – 1.135 | 0.933 | 0.977 | 0.808 – 1.181 | 0.810 |
| IMD: 5-6 vs IMD: 1-2 | 0.891 | 0.843 – 0.943 | **<0.001** | 1.086 | 0.978 – 1.206 | 0.122 | 0.998 | 0.874 – 1.139 | 0.971 | 1.087 | 0.899 – 1.314 | 0.389 |
| IMD: 7-10 vs IMD: 1-2 | 0.846 | 0.800 – 0.895 | **<0.001** | 1.099 | 0.989 – 1.220 | 0.079 | 0.977 | 0.856 – 1.116 | 0.732 | 1.128 | 0.933 – 1.364 | 0.213 |
| NL1 vs pre-pandemic : IMD: 3-4 vs IMD: 1-2 | 1.043 | 0.956 – 1.137 | 0.342 | 1.076 | 0.916 – 1.265 | 0.372 | 1.056 | 0.861 – 1.296 | 0.600 | 0.976 | 0.729 – 1.308 | 0.871 |
| NL2 vs pre-pandemic : IMD: 3-4 vs IMD: 1-2 | 1.033 | 0.935 – 1.143 | 0.522 | 1.006 | 0.834 – 1.214 | 0.947 | 1.070 | 0.845 – 1.357 | 0.574 | 0.984 | 0.701 – 1.382 | 0.926 |
| NL3 vs pre-pandemic : IMD: 3-4 vs IMD: 1-2 | 1.021 | 0.941 – 1.108 | 0.617 | 0.981 | 0.843 – 1.142 | 0.806 | 1.021 | 0.843 – 1.237 | 0.831 | 0.978 | 0.743 – 1.287 | 0.872 |
| NL1 vs pre-pandemic : IMD: 5-6 vs IMD: 1-2 | 1.043 | 0.956 – 1.137 | 0.342 | 1.124 | 0.956 – 1.320 | 0.158 | 1.064 | 0.867 – 1.305 | 0.553 | 1.037 | 0.774 – 1.389 | 0.810 |
| NL2 vs pre-pandemic : IMD: 5-6 vs IMD: 1-2 | 1.048 | 0.948 – 1.159 | 0.360 | 1.029 | 0.853 – 1.241 | 0.765 | 1.022 | 0.806 – 1.295 | 0.857 | 1.000 | 0.712 – 1.404 | 0.999 |
| NL3 vs pre-pandemic : IMD: 5-6 vs IMD: 1-2 | 1.025 | 0.944 – 1.112 | 0.559 | 1.002 | 0.861 – 1.166 | 0.980 | 1.025 | 0.846 – 1.242 | 0.803 | 0.960 | 0.729 – 1.264 | 0.773 |
| NL1 vs pre-pandemic : IMD: 7-10 vs IMD: 1-2 | 1.034 | 0.949 – 1.128 | 0.444 | 1.159 | 0.986 – 1.363 | 0.073 | 1.088 | 0.887 – 1.334 | 0.419 | 0.889 | 0.663 – 1.191 | 0.430 |
| NL2 vs pre-pandemic : IMD: 7-10 vs IMD: 1-2 | 1.048 | 0.948 – 1.159 | 0.362 | 1.037 | 0.860 – 1.251 | 0.701 | 1.018 | 0.803 – 1.290 | 0.885 | 1.006 | 0.716 – 1.413 | 0.973 |
| NL3 vs pre-pandemic : IMD: 7-10 vs IMD: 1-2 | 1.019 | 0.939 – 1.106 | 0.648 | 0.969 | 0.833 – 1.128 | 0.686 | 0.971 | 0.801 – 1.177 | 0.764 | 0.923 | 0.701 – 1.216 | 0.570 |
| | **Primary Care (ctd)** | | | | | | **Secondary Care** | | | | | |
| | **HbA1c** | | | **Smoking Status** | | | **Planned Admissions** | | | **Unplanned Admissions** | | |

| | RR | 95% CI | p-value | RR | 95% CI | p-value | RR | 95% CI | p-value | RR | 95% CI | p-value |
|---|---|---|---|---|---|---|---|---|---|---|---|---|
| NL1 vs pre-pandemic | 0.142 | 0.115 – 0.175 | **<0.001** | 0.352 | 0.309 – 0.401 | **<0.001** | **0.566** | **0.516 - 0.621** | **<0.001** | **0.642** | **0.558 - 0.737** | **<0.001** |
| NL2 vs pre-pandemic | 0.794 | 0.622 – 1.013 | 0.064 | 0.734 | 0.632 – 0.853 | **<0.001** | 0.916 | 0.823 - 1.020 | 0.110 | 0.944 | 0.803 - 1.109 | 0.479 |
| NL3 vs pre-pandemic | 0.871 | 0.714 – 1.061 | 0.171 | 0.735 | 0.651 – 0.831 | **<0.001** | **0.894** | **0.820 - 0.976** | **0.012** | 0.945 | 0.829 - 1.077 | 0.393 |
| IMD: 3-4 vs IMD: 1-2 | 0.947 | 0.781 – 1.149 | 0.582 | 0.977 | 0.868 – 1.101 | 0.707 | **0.761** | **0.699 - 0.828** | **<0.001** | 0.654 | 0.576 - 0.743 | **<0.001** |
| IMD: 5-6 vs IMD: 1-2 | 1.038 | 0.856 – 1.259 | 0.705 | 0.978 | 0.868 – 1.101 | 0.713 | **0.799** | **0.734 - 0.870** | **<0.001** | **0.598** | **0.527 - 0.680** | **<0.001** |
| IMD: 7-10 vs IMD: 1-2 | 1.041 | 0.858 – 1.263 | 0.681 | 0.972 | 0.863 – 1.094 | 0.636 | **0.848** | **0.779 - 0.923** | **<0.001** | **0.643** | **0.566 - 0.731** | **<0.001** |
| NL1 vs pre-pandemic : IMD: 3-4 vs IMD: 1-2 | 1.000 | 0.743 – 1.347 | 0.999 | 0.958 | 0.798 – 1.150 | 0.646 | 0.988 | 0.867 - 1.126 | 0.858 | 1.097 | 0.902 - 1.335 | 0.351 |
| NL2 vs pre-pandemic : IMD: 3-4 vs IMD: 1-2 | 0.991 | 0.702 – 1.401 | 0.961 | 0.997 | 0.806 – 1.233 | 0.977 | 1.001 | 0.860 - 1.165 | 0.990 | 1.017 | 0.810 - 1.277 | 0.883 |
| NL3 vs pre-pandemic : IMD: 3-4 vs IMD: 1-2 | 0.991 | 0.749 – 1.311 | 0.949 | 0.97 | 0.816 – 1.152 | 0.725 | 0.997 | 0.881 - 1.127 | 0.959 | 1.061 | 0.882 - 1.276 | 0.525 |
| NL1 vs pre-pandemic : IMD: 5-6 vs IMD: 1-2 | 1.020 | 0.758 – 1.374 | 0.895 | 0.974 | 0.811 – 1.170 | 0.779 | 0.765 | 0.671 - 0.872 | **<0.001** | 1.098 | 0.902 - 1.336 | 0.348 |
| NL2 vs pre-pandemic : IMD: 5-6 vs IMD: 1-2 | 1.009 | 0.714 – 1.426 | 0.958 | 0.933 | 0.754 – 1.153 | 0.519 | 1.043 | 0.896 - 1.214 | 0.588 | 0.981 | 0.781 - 1.233 | 0.871 |
| NL3 vs pre-pandemic : IMD: 5-6 vs IMD: 1-2 | 0.960 | 0.726 – 1.270 | 0.776 | 0.948 | 0.798 – 1.126 | 0.541 | 1.077 | 0.952 - 1.218 | 0.235 | 1.247 | 1.037 - 1.500 | **0.020** |
| NL1 vs pre-pandemic : IMD: 7-10 vs IMD: 1-2 | 0.929 | 0.690 – 1.251 | 0.629 | 0.984 | 0.819 – 1.181 | 0.861 | 0.695 | 0.609 - 0.792 | **<0.001** | 0.898 | 0.738 - 1.093 | 0.280 |
| NL2 vs pre-pandemic : IMD: 7-10 vs IMD: 1-2 | 1.019 | 0.721 – 1.440 | 0.915 | 0.852 | 0.689 – 1.054 | 0.139 | 1.032 | 0.887 - 1.202 | 0.678 | 1.173 | 0.934 - 1.473 | 0.169 |
| NL3 vs pre-pandemic : IMD: 7-10 vs IMD: 1-2 | 0.927 | 0.700 – 1.226 | 0.595 | 0.836 | 0.704 – 0.994 | **0.042** | 1.068 | 0.944 - 1.208 | 0.291 | 1.164 | 0.968 - 1.401 | 0.105 |

**Supplementary Table S10**: Associated interaction between the first national lockdown and deprivation group on primary HCU compared to pre-pandemic rate ratios between deprivation groups, for multi-morbid patients.

| | Deprivation 3-4 vs 1-2 | | Deprivation 5-6 vs 1-2 | | Deprivation 7-10 vs 1-2 | |
|---|---|---|---|---|---|---|
| | RR (95% CI) | p-value | RR (95% CI) | p-value | RR (95% CI) | p-value |
| **Primary HCU** | | | | | | |
| New prescriptions | 1.022 (0.913 – 1.145) | 0.697 | 1.012 (0.904 – 1.133) | 0.831 | 0.992 (0.886 – 1.111) | 0.888 |
| BP | 1.097 (0.886 – 1.358) | 0.390 | 1.127 (0.910 – 1.396) | 0.267 | 1.166 (0.942 – 1.444) | 0.156 |
| BMI | 1.018 (0.760 – 1.362) | 0.905 | 1.025 (0.766 – 1.372) | 0.865 | 1.102 (0.823 – 1.475) | 0.508 |
| Cholesterol | 0.958 (0.644 – 1.424) | 0.829 | 1.004 (0.675 – 1.492) | 0.986 | 0.853 (0.574 – 1.269) | 0.428 |
| HbA1c | 0.988 (0.665 – 1.467) | 0.950 | 0.996 (0.670 – 1.479) | 0.983 | 0.890 (0.599 – 1.323) | 0.561 |
| Smoking status | 0.939 (0.726 – 1.215) | 0.628 | 0.945 (0.731 – 1.223) | 0.665 | 0.957 (0.739 – 1.238) | 0.733 |
| **Secondary HCU** | | | | | | |
| Planned admissions | 1.011 (0.576 – 1.770) | 0.970 | 0.748 (0.392 – 1.404) | 0.371 | 0.731 (0.387 – 1.359) | 0.326 |
| Unplanned admissions | 1.153 (0.513 – 2.592) | 0.729 | 1.202 (0.490 – 2.919) | 0.685 | 0.889 (0.346 – 2.217) | 0.803 |

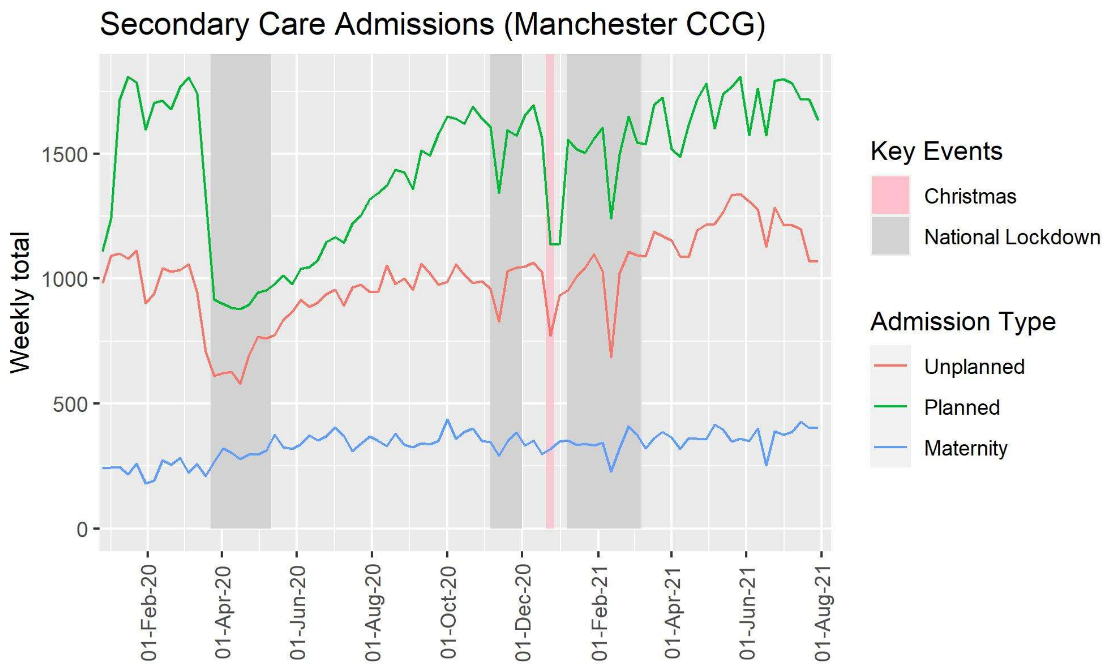


**Supplementary Figure S4**: Secondary healthcare utilisation across Manchester CCG, from January 2020 until August 2021.

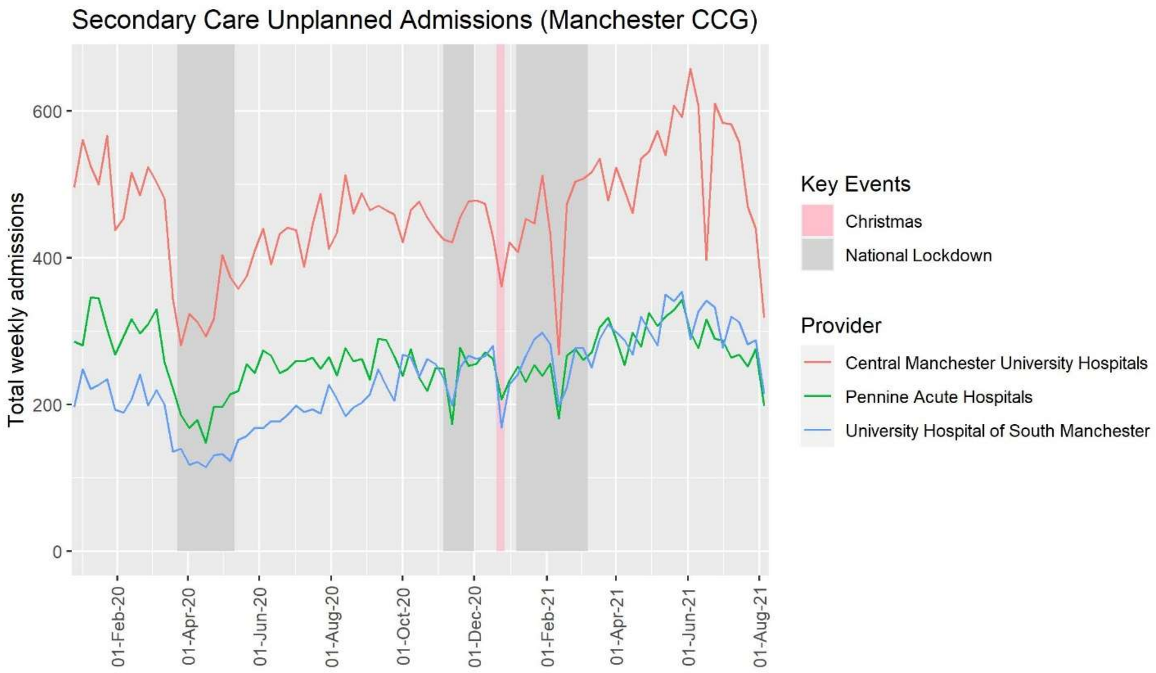


**Supplementary Figure S5:** Total weekly unplanned admissions of MCCG population at the three main providers (local to 96.6% of people) between January 2020 and August 2021.

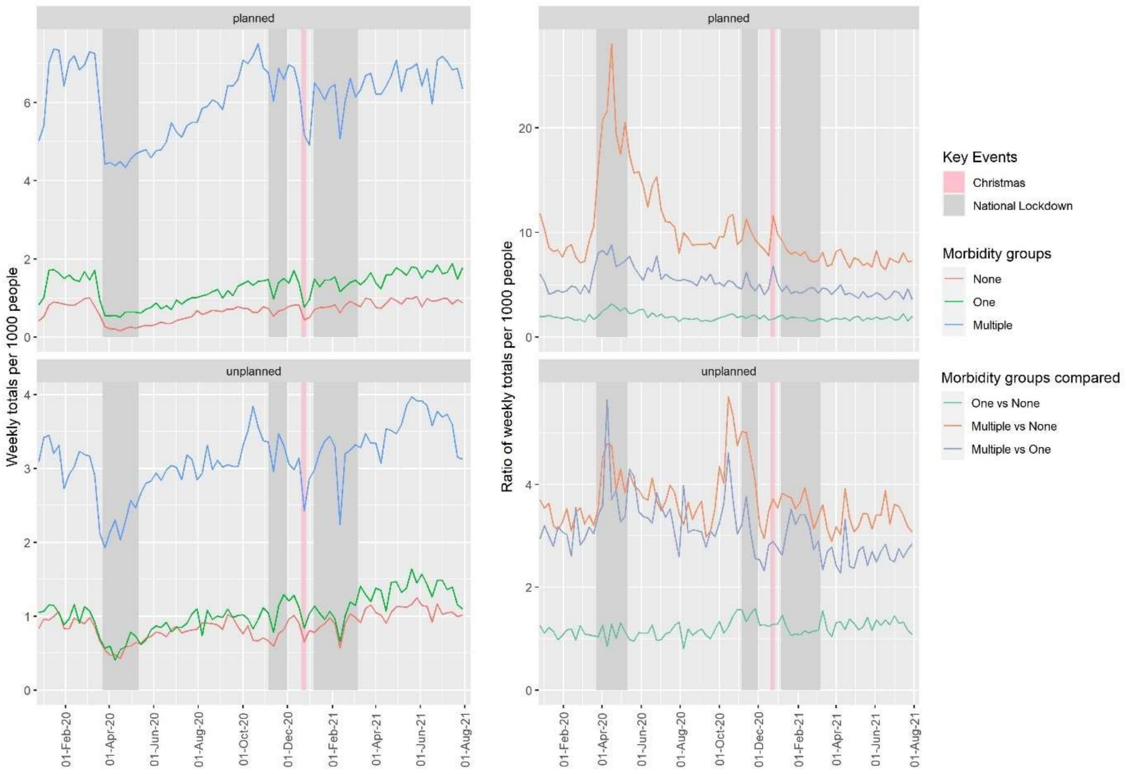


**Supplementary Figure S6:** Weekly planned and unplanned admissions per 1000 people within morbidity group in the MCCG population and the ratio between these scaled rates, between January 2020 and August 2021.

**Supplementary Table S11:** Estimates of the ratios of planned and unplanned admission rates for each LTC group in the final four weeks of the study compared to pre-pandemic rates.

| | Planned | | | Unplanned | | |
|---|---|---|---|---|---|---|
| LTC | Rate Ratio | 95% CI | p-value | Rate Ratio | 95% CI | p-value |
| Cancer | 0.653 | 0.586 – 0.726 | **<0.001** | 0.926 | 0.773 – 1.105 | 0.401 |
| Cardiovascular | 0.938 | 0.901 – 0.977 | **0.002** | 0.978 | 0.915 – 1.044 | 0.502 |
| Endocrine | 0.997 | 0.951 – 1.044 | 0.893 | 1.000 | 0.930 – 1.076 | 0.990 |
| Gastrointestinal | 0.959 | 0.921 – 0.998 | **0.039** | 1.104 | 1.039 – 1.171 | **0.001** |
| Musculoskeletal or Skin | 1.025 | 0.974 – 1.078 | 0.340 | 1.093 | 1.017 – 1.173 | **0.014** |
| Neurological | 1.053 | 0.908 – 1.218 | 0.489 | 1.137 | 0.946 – 1.360 | 0.165 |
| Psychiatric | 1.006 | 0.959 – 1.054 | 0.814 | 1.113 | 1.049 – 1.181 | **<0.001** |
| Renal or Urological | 0.926 | 0.885 – 0.969 | **0.001** | 0.839 | 0.750 – 0.938 | **0.002** |
| Respiratory | 0.966 | 0.910 – 1.025 | 0.255 | 1.020 | 0.947 – 1.098 | 0.597 |
| Sensory Impairment or Learning Disability | 0.894 | 0.841 – 0.950 | **<0.001** | 1.016 | 0.926 – 1.113 | 0.736 |
| Substance Abuse | 1.196 | 1.074 – 1.329 | **0.001** | 1.139 | 1.013 – 1.278 | **0.028** |

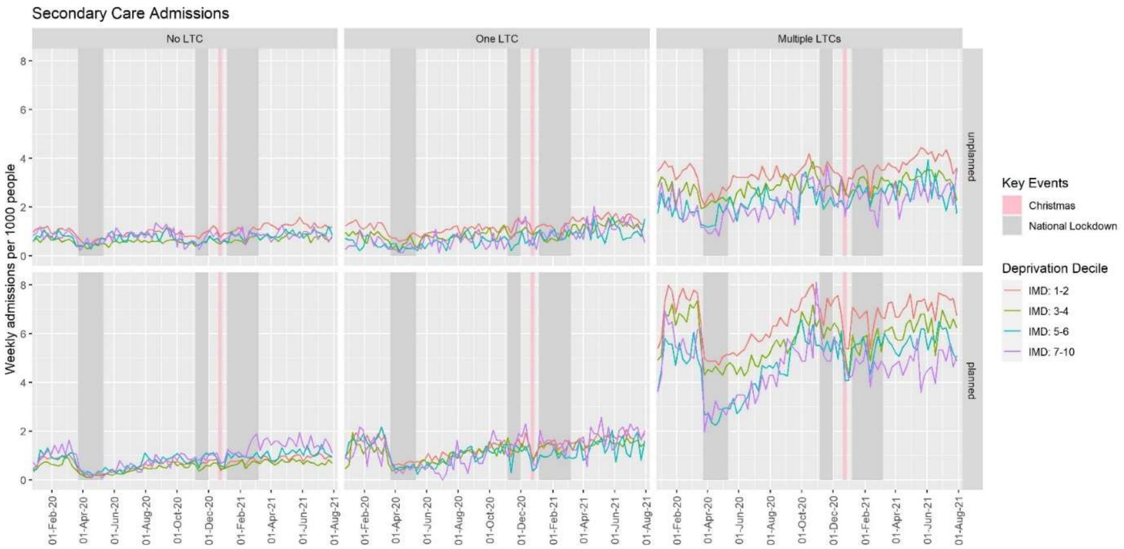


**Supplementary Figure S7:** Rates of weekly planned and unplanned admissions by deprivation and number of long-term conditions of the Manchester CCG population, between January 2020 and August 2021.

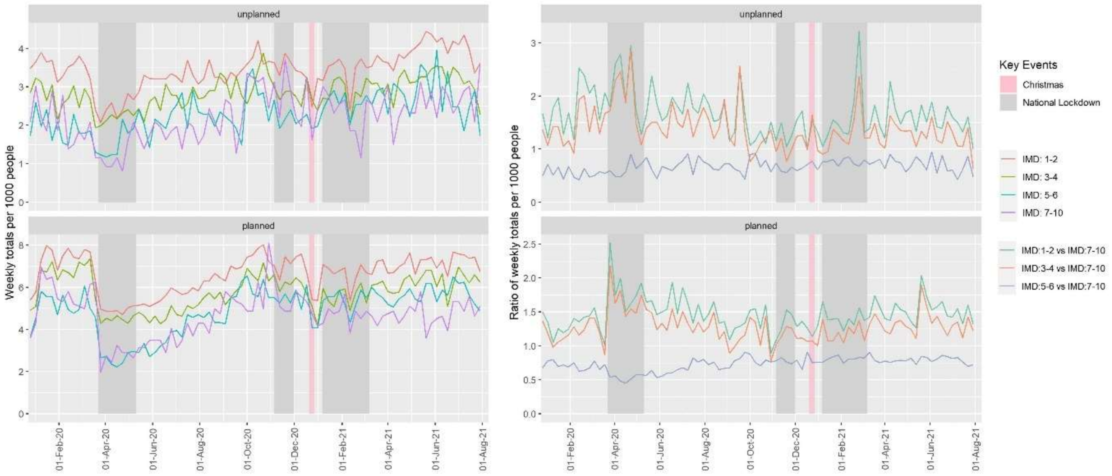


**Supplementary Figure S8:** Weekly planned and unplanned secondary care admissions per 1000 people within multi-morbid patients by deprivation group and the ratio of these compared to the least-deprived group (IMD: 7-10), between January 2020 and August 2021.

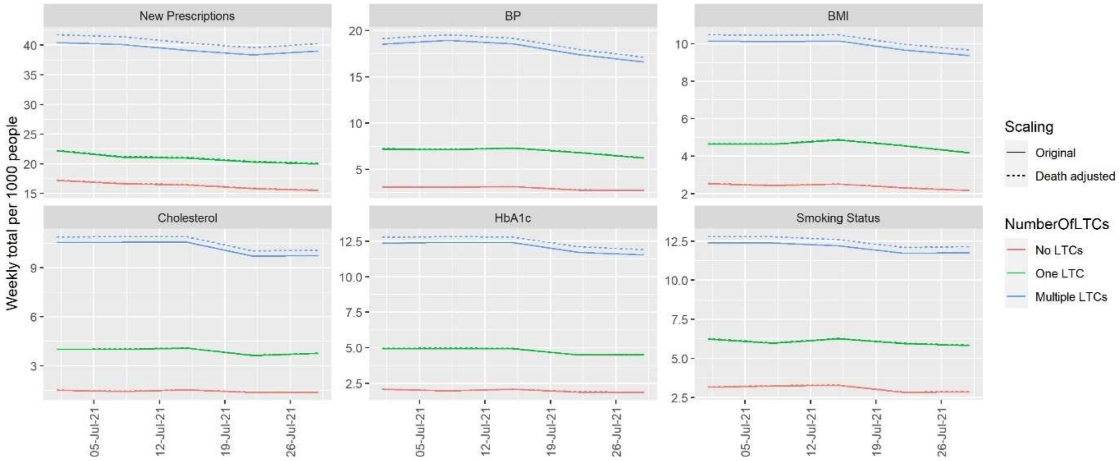


**Supplementary Figure S9:** Comparison between unadjusted (original) and death-adjusted weekly primary HCU measures per 1000 people, by morbidity in July 2021.

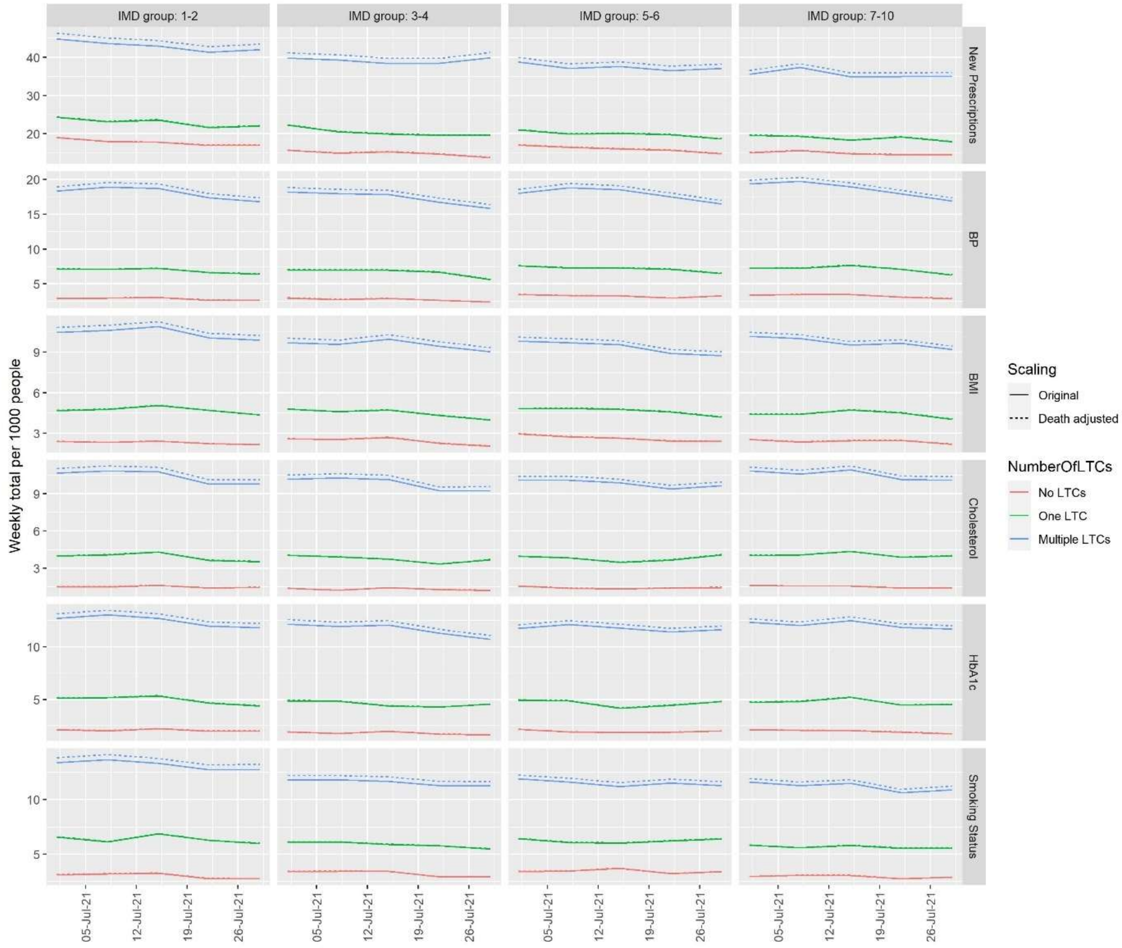


**Supplementary Figure 10:** Comparison between unadjusted (original) and death-adjusted weekly primary HCU measures per 1000 people, by deprivation group in July 2021.

**Supplementary Table S12:** Comparison between unadjusted (original) and death-adjusted weekly secondary care admissions per 1000 people, by morbidity group in July 2021.

| Number of LTCs (morbidity group) | Admission Type | Week start | Unadjusted Weekly admissions per 1000 people | Death-Adjusted Weekly admissions per 1000 people | Difference |
|---|---|---|---|---|---|
| None | unplanned | 01/07/2021 | 1.0235 | 1.0258 | 0.0023 |
| None | unplanned | 08/07/2021 | 1.0460 | 1.0483 | 0.0024 |
| None | unplanned | 15/07/2021 | 1.0560 | 1.0584 | 0.0024 |
| None | unplanned | 22/07/2021 | 0.9934 | 0.9957 | 0.0022 |
| None | unplanned | 29/07/2021 | 1.0185 | 1.0208 | 0.0023 |
| One | unplanned | 01/07/2021 | 1.4832 | 1.4898 | 0.0066 |
| One | unplanned | 08/07/2021 | 1.3576 | 1.3637 | 0.0060 |
| One | unplanned | 15/07/2021 | 1.3969 | 1.4031 | 0.0062 |
| One | unplanned | 22/07/2021 | 1.1536 | 1.1587 | 0.0051 |
| One | unplanned | 29/07/2021 | 1.0987 | 1.1035 | 0.0049 |
| Multiple | unplanned | 01/07/2021 | 3.6952 | 3.8118 | 0.1166 |
| Multiple | unplanned | 08/07/2021 | 3.7372 | 3.8551 | 0.1179 |
| Multiple | unplanned | 15/07/2021 | 3.5932 | 3.7066 | 0.1134 |
| Multiple | unplanned | 22/07/2021 | 3.1553 | 3.2548 | 0.0995 |
| Multiple | unplanned | 29/07/2021 | 3.1253 | 3.2239 | 0.0986 |
| None | planned | 01/07/2021 | 0.9859 | 0.9882 | 0.0022 |
| None | planned | 08/07/2021 | 0.9884 | 0.9907 | 0.0022 |
| None | planned | 15/07/2021 | 0.8458 | 0.8477 | 0.0019 |
| None | planned | 22/07/2021 | 0.9559 | 0.9581 | 0.0021 |
| None | planned | 29/07/2021 | 0.8733 | 0.8753 | 0.0020 |
| One | planned | 01/07/2021 | 1.6244 | 1.6317 | 0.0072 |

| | | | | | |
|---|---|---|---|---|---|
| One | planned | 08/07/2021 | 1.6558 | 1.6632 | 0.0073 |
| One | planned | 15/07/2021 | 1.8834 | 1.8918 | 0.0084 |
| One | planned | 22/07/2021 | 1.4832 | 1.4898 | 0.0066 |
| One | planned | 29/07/2021 | 1.7736 | 1.7814 | 0.0079 |
| Multiple | planned | 01/07/2021 | 7.1864 | 7.4131 | 0.2267 |
| Multiple | planned | 08/07/2021 | 7.0485 | 7.2708 | 0.2224 |
| Multiple | planned | 15/07/2021 | 6.8385 | 7.0542 | 0.2157 |
| Multiple | planned | 22/07/2021 | 6.8745 | 7.0914 | 0.2169 |
| Multiple | planned | 29/07/2021 | 6.3526 | 6.5530 | 0.2004 |
